# Treatment decision algorithms for tuberculosis screening and diagnosis in children below 5 years hospitalised with severe acute malnutrition: a cost-effectiveness analysis

**DOI:** 10.1101/2024.11.12.24317217

**Authors:** Marc d’Elbée, Nyashadzaishe Mafirakureva, Chishala Chabala, Minh Huyen Ton Nu Nguyet, Martin Harker, Clémentine Roucher, Gerald Businge, Perfect Shankalala, Bwendo Nduna, Veronica Mulenga, Maryline Bonnet, Eric Wobudeya, Olivier Marcy, Peter J. Dodd, TB-Speed SAM study group

**Affiliations:** University of Bordeaux, National Institute for Health and Medical Research (Inserm) UMR 1219, Research Institute for Sustainable Development (IRD) EMR 271, Bordeaux, France; Sheffield Centre for Health & Related Research (SCHARR), University of Sheffield, Sheffield, United Kingdom; School of Medicine, University of Zambia, Lusaka, Zambia; University Teaching Hospitals-Children’s Hospital, Lusaka, Zambia; TB Modelling Group, TB Centre, and Global Centre for Health Economics, London School of Hygiene and Tropical Medicine, London, United Kingdom; Makerere University-John Hopkins University Research Collaboration, Kampala, Uganda; Arthur Davidson Children’s Hospital, Ndola, Zambia; TransVIHMI, University of Montpellier, IRD, Inserm, Montpellier, France

**Keywords:** paediatric tuberculosis, severe acute malnutrition, treatment decision algorithms, diagnosis, economic evaluation, low- and middle-income countries, cost-effectiveness analysis

## Abstract

**Background:** Children with severe acute malnutrition (SAM) are an important risk group for underdiagnosis and death from tuberculosis. In 2022, the World Health Organization (WHO) recommended use of treatment decision algorithms (TDAs) for tuberculosis diagnosis in children. There is currently no cost-effectiveness evidence for TDA-based approaches compared to routine practice.

**Methods:** The TB-Speed SAM study developed i) a one-step TDA including Xpert, clinical, radiological and echography features, and ii) a two-step TDA, which also included a screening phase, for children under 5 years hospitalised with SAM at tertiary hospitals in Uganda and Zambia. We assessed the diagnostic accuracy and cost-effectiveness of deploying TB-Speed and WHO TDA-based approaches compared to the standard of care (SOC). Estimated outcomes included children started on tuberculosis treatment, false positive rates, disability-adjusted life years (DALYs) and incremental cost-effectiveness ratios (ICERs).

**Findings:** Per 100 children hospitalised with SAM, averaging 19 children with tuberculosis, the one-step TDA initiated 17 true positive children (95% uncertainty intervals [UI]: 12-23) on tuberculosis treatment, the two-step TDA 15 (95%UI: 10-22), the WHO TDA 14 (95%UI: 9-19), and SOC 4 (95%UI: 2-9). The WHO TDA generated the most false positives (35, 95%UI: 24-46), followed by the one-step TDA (18, 95%UI: 6-29), the two-step TDA (14, 95%UI: 1-25), and SOC (11, 95%UI: 3-17). All TDA-based approaches had ICERs below plausible country cost-effectiveness thresholds compared to SOC (one-step: $44-51/DALY averted, two-step: $34-39/DALY averted, WHO: $40-46/DALY averted).

**Interpretation:** Our findings show that these TDA-based approaches are highly cost-effective for the vulnerable group of children hospitalised with SAM, compared to current practice.

**Funding:** Unitaid

**Research in context:** *Evidence before this study:* In 2022, the WHO conditionally recommended the use of treatment decision algorithms (TDAs) for tuberculosis diagnosis in children aged <10 with presumptive pulmonary tuberculosis. Two TDAs were suggested for use in settings with (TDA A) and without (TDA B) access to chest X-ray. These WHO-suggested TDAs propose a single approach to TB diagnosis in all children. The TB Speed SAM study developed specific algorithms for children <5 years hospitalised with severe acute malnutrition. Aiming to identify studies assessing the cost-effectiveness of using TDAs for childhood TB, we searched the PubMed database using (“Decision Support Systems, Clinical”[MeSH] OR “clinical decision support” OR “decision support” OR “clinical decision-making”) AND (“Algorithms”[MeSH] OR “algorithm” OR “decision-making” OR “decision model” OR “treatment decision algorithm”) AND (“Tuberculosis”[MeSH] OR “tuberculosis” OR “TB”) AND (“Costs and Cost Analysis”[MeSH] OR “cost-effectiveness” OR “cost analysis” OR “costs”) between 1st January 2004 and 18th October 2024, without language restrictions. Of 31 articles identified, 2 articles reported on the cost-effectiveness of interventions aiming to improve clinical decision making for tuberculosis diagnosis. Other articles were excluded because they were not an economic evaluation, not on tuberculosis, or only compared microbiological testing approaches related to tuberculosis care (microscopic observation drug susceptibility test versus Xpert MTB/RIF test, QuantiFERON-TB Gold In-Tube versus tuberculin skin test for tuberculosis diagnosis). Debes et al. assessed the cost-effectiveness of tuberculosis diagnosis using microscopic observation drug susceptibility, Xpert MTB/RIF and empiric treatment for all patients, in addition to current clinical diagnostic practices in Ugandan children. Van’t Hoog et al. explored combinations of sensitivity, specificity and cost at which a hypothetical triage test would improve affordability of the Xpert assay. We found no economic evaluations of a treatment decision algorithm (TDA)-based approach (screening, testing, treatment) for tuberculosis diagnosis.

*Added value of this study:* This is the first study to assess the cost-effectiveness of using treatment decision algorithms in childhood tuberculosis diagnosis. It focuses on children <5 years hospitalised with severe acute malnutrition using the TB-Speed SAM one-step TDA and two-step TDA, which includes a screening step before the diagnostic step, and the WHO-suggested TDA A. We also evaluated the accuracy of the WHO-suggested TDA for paediatric tuberculosis in this patient group. This study found that for children hospitalised with SAM all three TDA-based approaches for paediatric tuberculosis diagnosis were cost-effective compared to the standard of care from a health systems perspective in Uganda and Zambia, including in lower tuberculosis prevalence settings. The TB-Speed two-step and WHO TDAs had lower costs than the TB-Speed one-step because their screening step resulted in fewer assessments overall, but also a smaller health impact due to a slightly lower sensitivity. The WHO TDA was less effective and more costly than the TB-Speed two-step TDA and involved substantial rates of overtreatment. The TB-Speed one-step TDA had the greatest health impact while remaining cost-effective, making it the preferred option.

*Implications of all the available evidence:* The WHO has conditionally recommended incorporating TDAs, pending validation, into existing case detection strategies to support the decentralisation of clinical tools and improve the identification of tuberculosis in children. Our findings show that TDA-based approaches are cost-effective for the vulnerable group of children hospitalised with SAM, compared to current practices, and our sensitivity analysis suggests that these results are robust. While not developed in children hospitalised with SAM, the WHO-suggested TDA for paediatric tuberculosis performs well in this patient group. This analysis contributes valuable evidence to support the interim WHO recommendation on decentralised models of care.

## Introduction

Tuberculosis mortality remains high in children globally, with 187,500 deaths estimated by the World Health Organization (WHO) for 1·25 million paediatric cases in 2023.^1,2^ The vast majority of childhood tuberculosis deaths occur because the disease remains undiagnosed and, therefore, untreated.^3^ Bacteriological tuberculosis diagnosis is challenging in children due to paucibacillary disease and challenging sample collection.^4^ Current diagnosis relies mostly on clinical and radiographic features, which lack specificity, notably in children with immunodeficiency (Vonasek et al. 2022). Recent studies show that children under 5 years of age with severe acute malnutrition (SAM) are an important risk group for the development of tuberculosis, and are at high risk of going undiagnosed.^5–7^

SAM is defined by WHO as severe wasting, i.e. low weight for height ratio, upper arm circumference (<115 mm), or clinical signs of bilateral nutritional pitting oedema,^8^ and is associated with high mortality. Childhood malnutrition is a major global health challenge with 45 million children affected, accounting for almost 50% of the deaths in children under the age of 5 years.^9,10^ The WHO estimates that undernutrition causes 20% of global tuberculosis cases, though the burden in children with SAM remains unclear (WHO 2024). Children with SAM require a different approach to TB diagnosis due to overlapping symptoms, non-specific radiological signs, poor test performance, and delayed diagnosis, increasing the risk of under-detection, mortality, and poor treatment outcomes (Vonasek et al. 2022). Therefore, improving case detection in children with SAM could contribute to reducing mortality in this vulnerable group and is essential to achieving the global target of zero deaths from tuberculosis in children by 2030.^6,7^

Treatment decision algorithms (TDAs) are scoring systems for clinical, radiological and microbiological features. Treatment initiation is recommended above a predetermined score threshold.^11^ Aiming to standardise and accelerate the identification of tuberculosis in children, TDAs could fill the detection gaps in vulnerable groups, often difficult to diagnose. WHO recently conditionally recommended the use of TDAs for the diagnosis of tuberculosis in children below 10 years old with symptoms suggestive of pulmonary tuberculosis.^12,13^ Specifically, WHO suggested using two TDAs for settings with chest X-ray [CXR] (algorithm A) and for settings without CXR (algorithm B). These two TDAs use the same diagnostic approach for the general paediatric population and highly vulnerable children such as those with severe acute malnutrition that may require customised approaches.

In 2023, the TB-Speed SAM study was the first to develop TDAs specifically for children under 5 years hospitalised with SAM at tertiary hospitals in Uganda and Zambia.^14^ Two TDAs were developed: i) a one-step TDA, including Xpert, clinical, radiological and echography features assessment in all children, and ii) a two-step TDA including a screening phase followed by similar assessment in those who screen positive. Chabala et al. found that both TDAs demonstrated satisfactory diagnostic accuracy with a sensitivity of 86.1% (95% confidence interval [CI]: 78.1–91.6) and specificity of 80.9% (95% CI: 76.9–84.3) for the one-step TDA, and a sensitivity and specificity of 79.2% (95% CI: 70.3–86.0) and 83.6% (95% CI: 79.9–86.8) for the two-step TDA, with a 30% reduction of tuberculosis assessments needed, due to the screening step^14^.

In this study, we sought to assess the overall diagnostic accuracy and quantify the cost-effectiveness of the two TB-Speed SAM TDAs and the WHO-suggested TDA A (with CXR; WHO TDA hereafter) compared to routine clinical practice, from a health system perspective, for tuberculosis screening and diagnosis in children hospitalised with SAM.

## Methods

### Study design

We conducted a cost-effectiveness analysis of the TB-Speed SAM study using patient pathways, costs and cost-effectiveness modelling. We assessed the two TB-Speed SAM TDAs and the WHO TDA compared to routine clinical practice, from a health system perspective. The TB-Speed SAM study design has been described elsewhere.^14^ In addition, we evaluated the diagnostic accuracy of the WHO TDA.

The study was approved by the sponsor’s institutional review committee, the WHO ethical review committee, and the national ethics committees and institutional review boards in Uganda and Zambia. Written informed consent was obtained from parents or guardians. The TB-Speed SAM study is registered with clinicaltrials.gov (NCT04240990).

### Patient pathways

Approximately 10 international and local medical doctors, who are engaged in both clinical practice and research at tertiary-level hospitals, contributed expert input for the modelling assumptions used in the study. We developed conceptual models with country experts to represent patient care pathways and resource use for four comparator arms: a standard of care (SOC), the one-step TDA, two-step TDA, and the WHO TDA (**Figure 1**). The SOC pathway represented typical care in tertiary level hospitals in Uganda and Zambia, with a mix of options for assessment. The one-step and two-step approaches were based on the TB-Speed SAM study protocol and results,^14^ and the WHO TDA was based on the 2022 WHO operational handbook.^12^ (**Appendix figures 1, 2** and **3**). The TB-Speed SAM one-step TDA proposes systematic Xpert testing on stool and gastric aspirate samples, clinical evaluation, CXR and abdominal ultrasound echography. Children with a score ≥10 during clinical assessment are immediately initiated on TB treatment and do not receive echography. They receive Xpert testing for bacteriological confirmation and to assess whether there is drug-resistance as well as CXR to assess the disease severity. The two-step TDA presents a screening phase of clinical examination and HIV testing, then is similar to the one-step TDA for those with presumptive tuberculosis. The WHO TDA uses similar features to the one-step TDA, except for echography, and in that it is used in children with presumed tuberculosis on the basis of chronic symptoms.

**Figure 1.**
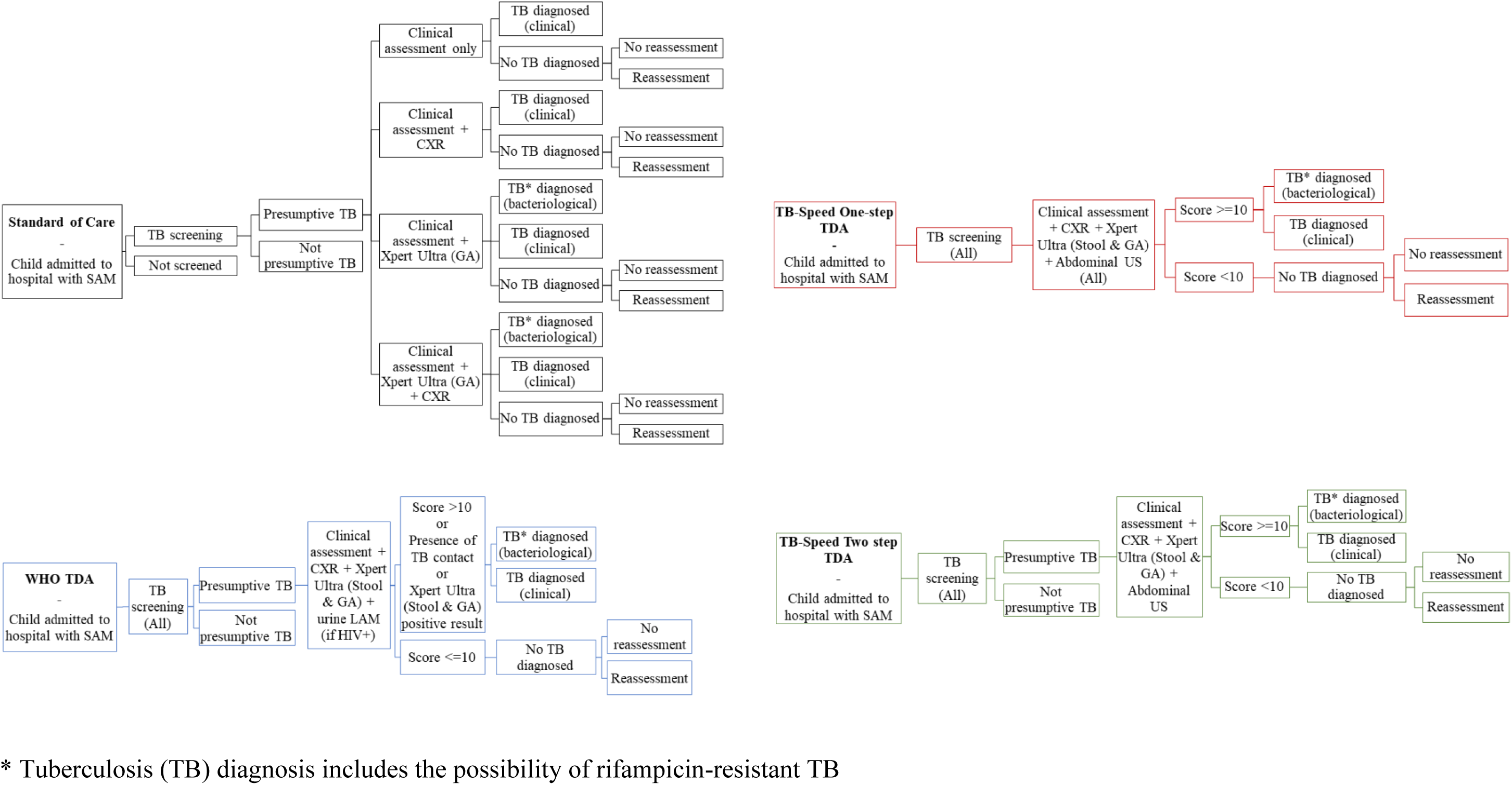
Model patient care pathways for the diagnosis and treatment of tuberculosis in children hospitalised with severe acute malnutrition (SAM). CXR = chest X-ray; GA = gastric aspirate; LAM = lipoarabinomannan lateral flow assay; TB = tuberculosis; TDA = treatment decision algorithm

**Figure 2.**
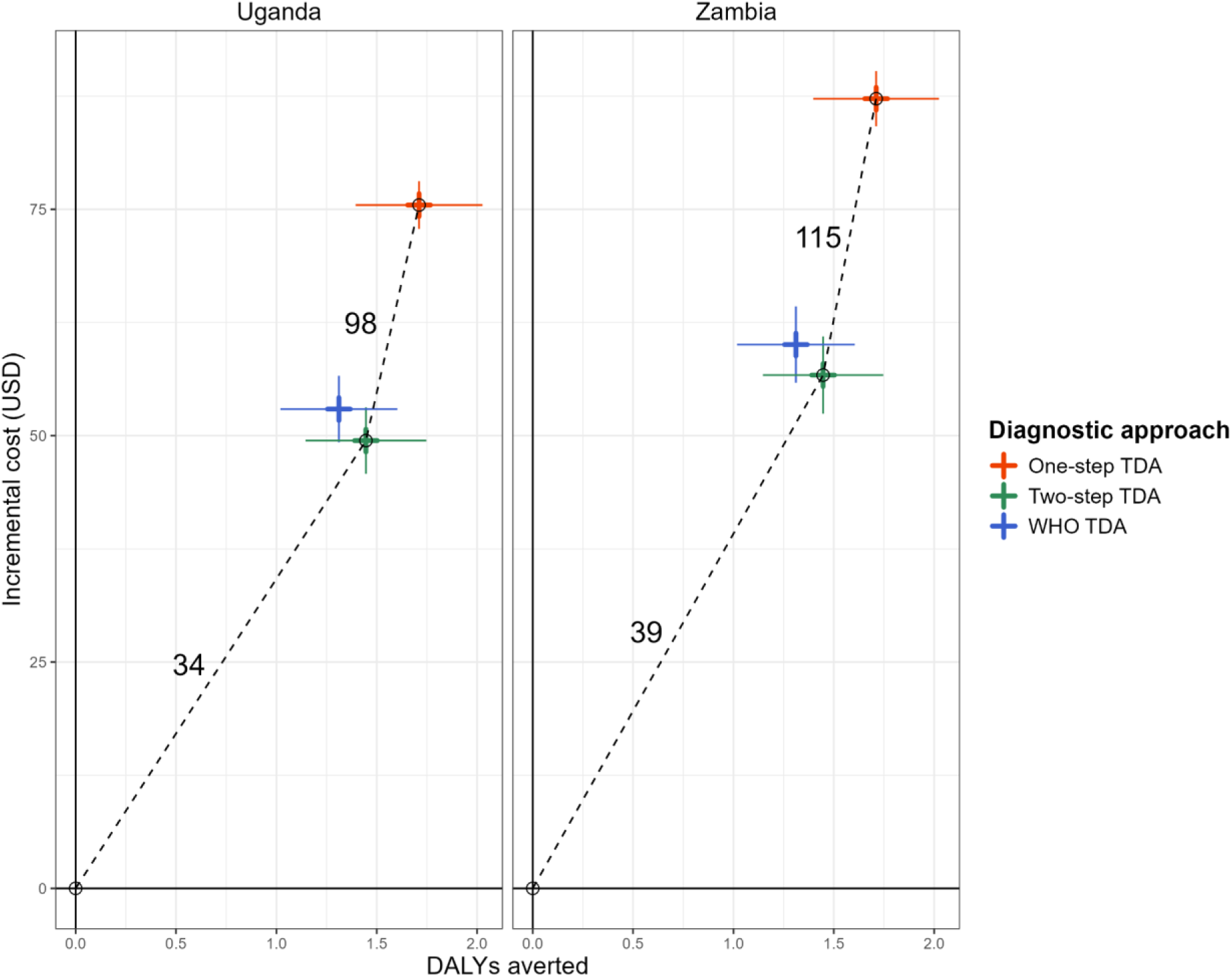
Cost-effectiveness planes for the TB-Speed SAM and WHO treatment decision algorithms, compared to the standard of care, by country. DALYs: disability-adjusted life years. Numerical annotations represent pairwise incremental cost-effectiveness ratios corresponding to the dashed line of the convex hull. Error bars represent one standard deviation.

**Figure 3.**
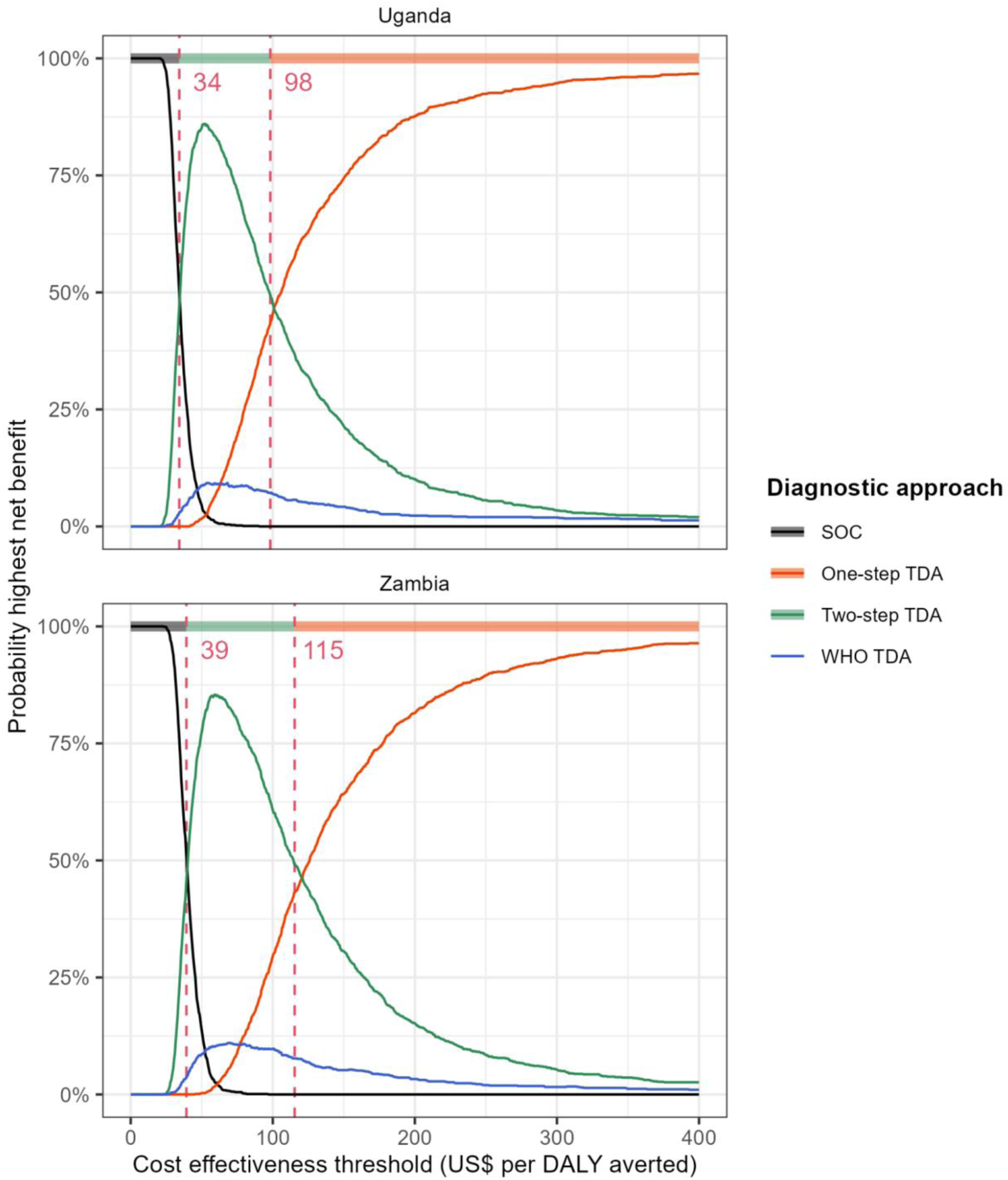
Cost-effectiveness acceptability frontiers and probability of highest net benefit for the TB-Speed SAM and WHO treatment decision algorithms and the standard of care, by country (in US$ per DALY averted). DALYs: disability-adjusted life years. Lines represent the proportion of simulations in which each option had the highest net benefit. Colours along the top denote the optimal choice (the option with the highest mean net benefit across all simulations) at each threshold. Dashed lines show the values of the cost-effectiveness threshold at which the optimal choice changes.

We based the diagnostic accuracy of screening for initial assessment and reassessment on data from TB-Speed SAM.

### Screening for tuberculosis

In the SOC, children admitted to hospital with SAM received non-systematic screening for tuberculosis, with a coverage of 80% suggested by clinicians. For the TDA arms, systematic screening for tuberculosis was conducted for all. Children were assumed to screen positive if they exhibited any respiratory symptoms, clinical signs, or other risk factors, such as HIV or a history of tuberculosis contact. We did not consider tuberculosis rescreening for any arm. See **Appendix table 1** and **2** for details.

### Reassessment following negative initial tuberculosis assessment

Using the TB-Speed SAM data, we defined a reassessment process for all arms, using i) the sensitivity and specificity of the clinician’s choice to reassess; and ii) the sensitivity and specificity of the reassessment examination. Reassessment was taken to comprise CXR, Xpert on gastric aspirate, and clinical assessment, and their sensitivity and specificity were conditional on results of the initial assessment being negative (**Appendix section 4**). We assumed that children can only be reassessed once.

### Evaluation of TDA diagnostic accuracy

A synthetic population of 10,000 children was created matching frequency cross-tabulations of signs and symptoms among children evaluated as true tuberculosis and not tuberculosis by an expert committee within the TB-Speed SAM study (N=535)^14^, and scores calculated for each TDA. Tuberculosis screening variables were resampled from the TB-Speed SAM cohort to account for higher order correlations. Resampling was also conducted on all variables of the TB-Speed SAM cohort to ensure unbiased replication for our synthetic cohort and found acceptable (results not shown). To maintain full reproducibility while not including data allowing reconstruction of the individual cohort data, we used a hybrid approach of modelling the screening variables using resampled data, while retaining the synthetic cohort-based approach to other variables. Symptoms included fatigue, loss of appetite, fever, and weight loss. Signs included results from the clinical exam, radiological testing (CXR), abdominal ultrasound, and microbiological test results (Xpert and HIV testing). Two symptoms (night sweat and haemoptysis) used in the WHO diagnostic score were not collected in TB-Speed SAM; their tuberculosis-stratified frequency was based on Gunasekera and al.^13^ Diagnostic accuracy of all TDAs for the synthetic cohort was assessed against expert committee tuberculosis status from the TB-Speed SAM study, using the updated NIH Clinical Case Definition.^14,15^

### Costing approach

Cost data collection tools were adapted from the Value TB costing tool suite, with reference to the WHO guidelines.^16^ Labour costs were sourced from national pay scales and project accounts, medications from the Global Drug Facility catalogue^17^, consumables, staff training, and equipment from project accounts, and hospitalisation cost of an inpatient bed day from WHO-CHOICE unit cost estimates.^18^ As this intervention did not change facility infrastructure, we excluded facility-associated overhead costs from the analysis.

Proportionate use of major equipment such as X-ray or Xpert machines was based on expected lifespan and annual number of uses. Key informants at MSF-Logistique (https://www.msflogistique.org/) estimated expected lifespan and laboratory managers in the three tertiary hospitals provided the number of annual uses. To value the contribution of labour, we conducted a time and motion study to estimate the length of time that staff spent on each patient care task under the TB-Speed intervention.^19^

Unit costs were estimated using ingredient-based costing. Services provided to patients were valued by multiplying quantities required by their unit costs. Costs were estimated in 2021 US dollars (US$), using a discount rate of 3% to annualise equipment costs.^20,21^ Lateral flow urine lipoarabinomannan (LF-LAM) test costs were based on literature.^22^ Since all TB care costs were incurred within a single year, discounting was not required. See **Appendix table 5** and **6** for cost parameters.

### Modelling approach

A decision-analytic cost-effectiveness model was developed in R software (version 4·3·0) to estimate the clinical benefits and cost-effectiveness of each diagnostic approach. A patient-level decision tree model represented clinical pathways shown in **Figure 1**, with outcomes depending on true tuberculosis status. The probabilities of having tuberculosis, diagnosis, treatment, and death were based on TB-Speed SAM study data and expert opinion. In particular, counts of children among the cohort stratified by tuberculosis status were used to parametrize beta distributions for branching probabilities, assuming uniform priors. See **Appendix table 7** for non-cost parameters.

### Outcomes

For each arm, we report the sensitivity and specificity of the tuberculosis screening step only, and of the overall arm (screening, initial assessment and reassessment), false positive and false negative rates, positive predictive value (PPV), and negative predictive value (NPV). We computed country-specific total and incremental mean costs and life-years lost over a lifetime horizon (with and without 3% discounting). Case fatality rates by tuberculosis status and treatment status were estimated from the TB-Speed SAM cohort and used to calculate deaths averted between arms. We disregarded the contribution of morbidity to disability adjusted life-years (DALYs), which has previously been shown to be a good approximation.^23^ We calculated DALYs using the life expectancy tables provided by the United Nations. These metrics were reported per 100 children admitted to hospital with SAM.

We report incremental cost-effectiveness ratios (ICERs) for all diagnostic approaches in each country. An estimated cost-effectiveness thresholds range for each country was used to assess potential cost-effectiveness.^24^ We also report cost-effectiveness acceptability frontiers (CEAF).^25^ We present our findings at the cohort tuberculosis prevalence (18·9%) and at lower prevalence levels similar to those reported in other studies on children with SAM (10·0%, 5·0%).^5^ All results, including equal-tailed 95% uncertainty intervals, were calculated using a probabilistic sensitivity analysis using Monte Carlo simulations with 1,000 parameter and cohort samples.

A health economic analysis plan was developed and is available upon request. We complied with the Consolidated Health Economic Evaluation Reporting Standards (CHEERS 2022) reporting guidelines.^26^

### Sensitivity analysis

We conducted sensitivity analyses to assess the impact of all model parameters on estimated ICERs compared to the SOC, using the interquartile range’s lower and upper limits, differentiated by TDA approach and by country.

## Results

### Accuracy of the SOC and TDA-based approaches

The two-step TDA had the highest screening sensitivity (89%, 95% uncertainty interval (UI): 81-94) followed by the WHO TDA (80%, 95% UI: 71-87), whereas the SOC had the lowest sensitivity (37%, 95% UI: 28-48) explained by an assessment solely based on the presence of chronic fever or cough (>2 weeks), and tuberculosis contact history (**Table 1**). Screening specificity was the highest for SOC (79%, 95% UI: 75-83), followed by two-step TDA (35%, 95% UI: 30-39), and WHO TDA (28%, 95% UI: 23-32). Combined screening and treatment decision sensitivity was the highest for one-step TDA (92%, 95% UI: 85-98), followed by the two-step TDA (82%, 95% UI: 72-92), the WHO TDA (77%, 95% UI: 64-88) and the SOC (26%, 95% UI: 15-38). The overall specificity was the lowest for the WHO TDA (57%, 95% UI: 51-63), followed by the one-step TDA (78%, 95% UI: 73-83), the two-step TDA (84%, 95% UI: 79-88) and the SOC (87%, 95% UI: 83-91).

**Table 1.** Accuracy of the standard of care, TB-Speed SAM and WHO treatment decision algorithms at classifying tuberculosis in 100 children hospitalised with severe acute malnutrition (SAM) at varying tuberculosis prevalence.

| Diagnostic approach | Standar<br>d of<br>care | One-step<br>TDA | Two-step<br>TDA | WHO<br>TDA | Standar<br>d of<br>care | One-step<br>TDA | Two-step<br>TDA | WHO<br>TDA | Standar<br>d of<br>care | One-step<br>TDA | Two-step<br>TDA | WHO<br>TDA |
| --- | --- | --- | --- | --- | --- | --- | --- | --- | --- | --- | --- | --- |
| Tuberculosis prevalence (%) | 18·9 (17·7 - 20·0)<sup>a</sup> |  |  |  | 10·0 (9·0 - 11·0) |  |  |  | 5·0 (4·5 - 5·5) |  |  |  |
| Cascade of care (per 100 children hospitalised with SAM) – n (95% UI) |  |  |  |  |  |  |  |  |  |  |  |  |
| *Screened for tuberculosis* | 80 (70 - 90) | 100 (100 - 100) | 100 (100 - 100) | 100 (100 - 100) | 80 (70 - 90) | 100 (100 - 100) | 100 (100 - 100) | 100 (100 - 100) | 80 (70 - 90) | 100 (100 - 100) | 100 (100 - 100) | 100 (100 - 100) |
| *Assessed for tuberculosis* | 19 (15 - 24) | 100 (100 - 100) | 70 (64 - 75) | 74 (69 - 79) | 18 (14 - 22) | 100 (100 - 100) | 68 (62 - 73) | 73 (68 - 78) | 17 (13 - 22) | 100 (100 - 100) | 67 (61 - 72) | 73 (68 - 78) |
| *Reassessed for tuberculosis* | 2 (0 - 3) | 21 (16 - 25) | 14 (10 - 18) | 8 (6 - 11) | 1 (0 - 3) | 21 (17 - 26) | 14 (10 - 18) | 8 (5 - 11) | 1 (0 - 3) | 22 (17 - 26) | 14 (10 - 18) | 8 (6 - 12) |
| *Initiated on tuberculosis treatment* | 15 (12 - 19) | 35 (29 - 41) | 29 (23 - 35) | 49 (43 - 55) | 14 (10 - 18) | 29 (22 - 38) | 23 (16 - 31) | 46 (40 - 53) | 14 (10 - 18) | 26 (20 - 36) | 20 (14 - 29) | 45 (38 - 51) |
| *Initiated on tuberculosis treatment*<br>*(true positive children only)* | 4 (2 - 9) | 17 (12 - 23) | 15 (10 - 22) | 14 (9 - 19) | 2 (0 - 6) | 8 (5 - 14) | 8 (4 - 14) | 7 (4 - 11) | 1 (0 - 6) | 4 (2 - 8) | 4 (1 - 8) | 3 (1 - 6) |
| *Initiated on tuberculosis treatment*<br>*at initial assessment* | 15 (12 - 19) | 34 (28 - 41) | 28 (22 - 35) | 48 (42 - 55) | 14 (10 - 18) | 28 (21 - 38) | 23 (15 - 31) | 46 (39 - 53) | 14 (10 - 18) | 25 (19 - 36) | 20 (14 - 29) | 45 (37 - 51) |
| **Diagnostic accuracy metrics of arm – % (95% UI)** |  |  |  |  |  |  |  |  |  |  |  |  |
| *False Positive Rate* | 13 (9 - 17) | 22 (17 - 27) | 16 (12 - 21) | 43 (37 - 49) | 13 (9 - 17) | 22 (17 - 27) | 16 (12 - 21) | 43 (37 - 48) | 13 (9 - 17) | 22 (17 - 27) | 16 (12 - 21) | 43 (37 - 49) |
| *False Negative Rate* | 74 (62 - 85) | 8 (2 - 15) | 18 (8 - 28) | 23 (12 - 36) | 74 (54 - 93) | 8 (0 - 21) | 18 (0 - 36) | 24 (5 - 43) | 74 (0 - 100) | 8 (0 - 33) | 17 (0 - 54) | 24 (0 - 71) |
| *Positive Predictive Value* | 31 (17 - 49) | 49 (42 - 57) | 54 (44 - 64) | 29 (23 - 35) | 18 (4 - 36) | 31 (24 - 39) | 36 (25 - 48) | 16 (11 - 22) | 9 (0 - 36) | 18 (11 - 23) | 21 (10 - 30) | 8 (3 - 12) |
| *Negative Predictive Value* | 83 (80 - 86) | 97 (95 - 99) | 95 (92 - 97) | 91 (85 - 95) | 91 (88 - 93) | 98 (96 - 100) | 97 (95 - 100) | 95 (91 - 99) | 95 (94 - 100) | 99 (97 - 100) | 98 (96 - 100) | 97 (93 - 100) |
| *Screening sensitivity<sup>b</sup>* | 37 (28 - 48) | 100 (100 - 100) | 89 (81 - 94) | 80 (71 - 87) | 37 (28 - 48) | 100 (100 - 100) | 89 (81 - 94) | 80 (71 - 87) | 37 (28 - 48) | 100 (100 - 100) | 89 (81 - 94) | 80 (71 - 87) |
| *Screening specificity<sup>b</sup>* | 79 (75 - 83) | 0 (0 - 0) | 35 (30 - 39) | 28 (23 - 32) | 79 (75 - 83) | 0 (0 - 0) | 35 (30 - 39) | 28 (23 - 32) | 79 (75 - 83) | 0 (0 - 0) | 35 (30 - 39) | 28 (23 - 32) |
| *Overall arm sensitivity* | 26 (15 - 38) | 92 (85 - 98) | 82 (72 - 92) | 77 (64 - 88) | 26 (7 - 46) | 92 (79 - 100) | 82 (64 - 100) | 76 (57 - 95) | 26 (0 - 100) | 92 (67 - 100) | 83 (46 - 100) | 76 (29 - 100) |
| *Overall arm specificity* | 87 (83 - 91) | 78 (73 - 83) | 84 (79 - 88) | 57 (51 - 63) | 87 (83 - 91) | 78 (73 - 83) | 84 (79 - 88) | 57 (52 - 63) | 87 (83 - 91) | 78 (73 - 83) | 84 (79 - 88) | 57 (51 - 63) |

<sup>a</sup>Prevalence of tuberculosis in the TB-Speed SAM study, <sup>b</sup>Uncertainty intervals are exact binomial confidence intervals from the actual cohort, UI: uncertainty interval

Per 100 children hospitalised with SAM, tuberculosis diagnostic assessment was much higher in the three TDA arms (70 [95% UI: 64-75] to 100 [95% UI: 100-100] children) compared to the SOC (19 children, 95% UI: 15-24). The WHO TDA initiated the highest number of children on tuberculosis treatment (49, 95% UI: 43-55), followed by the one-step TDA (35, 95% UI: 29-41), the two-step TDA (29, 95% UI: 23-35), and only 15 (95% UI: 12-19) children were initiated on treatment in the SOC. However, as shown by the overall arm sensitivity (1 – false negative rate) and specificity (1 – false positive rate), the WHO TDA also presented the highest rate of false positives (43, 95% UI: 37-49), followed by the one-step TDA (22, 95% UI: 17-27), the two-step TDA (16, 95% UI: 12-21) and the SOC (13, 95% UI: 9-17). Therefore, the number of true positive cases initiated on treatment was similar across the three TDA arms (14 [95% UI: 9-19] to 17 [95% UI: 12-23] children) and was three to four times higher than the SOC (4 [95% UI: 2-9] children). Across all arms, the vast majority of children treated (>97%, 95% UI: 96-100) were initiated on tuberculosis treatment following the first assessment. PPV was highest in the two-step TDA (54%, 95% UI: 44-64) and was lowest in the WHO TDA (29%, 95% UI: 23-35), whereas NPV was highest in the one-step TDA (97%, 95% UI: 95-99) and lowest in the SOC (83%, 95% UI: 80-86). At lower tuberculosis prevalence, the NPV remained high, but the PPV significantly decreased due to the rarity of true positive children detected.

### Costs and cost-effectiveness

Costs were slightly higher in Zambia than in Uganda (**Table 2**). We present the results for Uganda, with the full model outputs (both discounted and undiscounted) provided in the appendix at varying levels of tuberculosis prevalence (**Appendix table 8**). Incremental costs per child going through the patient care pathway were highest for the one-step TDA ($75, 95% UI: 70-80), then for the WHO TDA ($53, 95% UI: 46-60), followed by the two-step TDA ($49, 95% UI: 42-57), compared to the SOC. Compared to the SOC, the three TDA-based approaches had similar impact on child deaths averted (5, 95% UI: 3-7), resulting in DALYs averted of 171 (95% UI: 113-235) for the one-step TDA, 145 (95% UI: 88-201) for the two-step TDA, and 131(95% UI: 78-190) for the WHO TDA.

**Table 2.** Costs and cost-effectiveness of the tuberculosis diagnostic approaches by country.

|  | Arm (versus Comparator) | Uganda | Zambia |
| --- | --- | --- | --- |
| **Per child hospitalised with SAM** |  |  |  |
| Costs per child (95% UI) in 2021 US dollars | SOC - Total | 32 (29 - 36) | 38 (33 - 42) |
|  | WHO TDA - Total | 85 (79 - 92) | 98 (90 - 105) |
|  | Two-step TDA - Total | 82 (75 - 88) | 94 (87 - 102) |
|  | One-step TDA - Total | 108 (104 - 112) | 125 (121 - 129) |
| Incremental costs per child (95% UI) in 2021 US dollars | WHO TDA (vs. SOC) | 53 (46 - 60) | 60 (52 - 68) |
|  | Two-step TDA (vs. SOC) | 49 (42 - 57) | 57 (48 - 65) |
|  | One-step TDA (vs. SOC) | 75 (70 - 80) | 87 (81 - 93) |
|  | Two-step TDA (vs WHO TDA) | -3 (-11 - 4) | -3 (-12 - 5) |
|  | One-step TDA (vs. WHO TDA) | 23 (16 - 29) | 27 (20 - 35) |
|  | One-step TDA (vs. Two-step TDA) | 26 (20 - 32) | 31 (24 - 37) |
| **Per 100 children hospitalised with SAM** |  |  |  |
| Deaths (95% UI) | SOC - Total | 19 (16 - 21) | 19 (16 - 21) |
|  | WHO TDA - Total | 14 (13 - 15) | 14 (13 - 15) |
|  | Two-step TDA - Total | 13 (12 - 15) | 13 (12 - 14) |
|  | One-step TDA - Total | 12 (12 - 13) | 12 (12 - 13) |
| Deaths averted (95% UI) | WHO TDA (vs. SOC) | 5 (3 - 7) | 5 (3 - 7) |
|  | Two-step TDA (vs. SOC) | 5 (3 - 7) | 5 (3 - 8) |
|  | One-step TDA (vs. SOC) | 6 (4 - 9) | 6 (4 - 8) |
|  | Two-step TDA (vs WHO TDA) | 0 (-1 - 2) | 0 (-1 - 2) |
|  | One-step TDA (vs. WHO TDA) | 1 (0 - 3) | 1 (0 - 3) |
|  | One-step TDA (vs. Two-step TDA) | 1 (0 - 2) | 1 (0 - 2) |
| Discounted DALYs (95% UI) | SOC - Total | 509 (449 - 570) | 509 (450 - 571) |
|  | WHO TDA - Total | 377 (347 - 414) | 378 (347 - 415) |
|  | Two-step TDA - Total | 364 (337 - 396) | 364 (337 - 394) |
|  | One-step TDA - Total | 338 (321 - 358) | 338 (321 - 359) |
| Incremental discounted<br>DALYs averted (95%<br>UI) | WHO TDA (vs. SOC) | 131 (78 - 190) | 131 (77 - 191) |
|  | Two-step TDA (vs. SOC) | 145 (88 - 201) | 145 (91 - 205) |
|  | One-step TDA (vs. SOC) | 171 (113 - 235) | 171 (113 - 232) |
|  | Two-step TDA (vs WHO TDA) | 13 (-20 - 49) | 14 (-20 - 48) |
|  | One-step TDA (vs. WHO TDA) | 40 (10 - 75) | 40 (11 - 76) |
|  | One-step TDA (vs. Two-step TDA) | 26 (7 - 51) | 26 (7 - 51) |
| Discounted ICER | WHO TDA (vs. SOC) | 40 | 46 |
|  | Two-step TDA (vs. SOC) | 34 | 39 |
|  | One-step TDA (vs. SOC) | 44 | 51 |
|  | Two-step TDA (vs WHO TDA) | Two-step TDA<br>dominates WHO TDA | Two-step TDA<br>dominates WHO TDA |
|  | One-step TDA (vs. Two-step TDA) | 98 | 115 |

SOC: standard of care, SAM: severe acute malnutrition, DALYs: disability-adjusted life years, ICER: incremental cost-effectiveness ratio, UI: uncertainty interval, TDA: treatment decision algorithm
Costs and life expectancy tables for estimating DALYs are specific to each country. Incremental costs compare costs of different approaches.

Incremental costs and DALYs averted are shown on a cost-effectiveness plane (**Figure 2**). All three TDA-based approaches compared to the SOC presented ICERs well below estimates of implied country cost-effectiveness thresholds (UG: 150-194 $/DALY averted, ZM: 364-500 $/DALY averted, **Appendix table 9**). The probability of two-step TDA being cost-effective at these thresholds compared to SOC was close to 100%, with ICERs of $34/DALY averted (UG) or $39/DALY averted (ZM) (**Figure 2**). The one-step TDA was more effective but costlier, whereas the WHO TDA was less effective but costlier than the two-step TDA. ICERs for the one-step TDA compared to the two-step TDA (UG: $98/DALY averted, ZM: $115/DALY averted) are also below the country-specific threshold ranges, making this highly likely to be cost-effective (**Figure 2**). The CEAF indicates that in both countries, the one-step TDA is the optimal choice, offering the highest mean net benefit and remaining cost-effective (**Figure 3**).

### Sensitivity analysis

The parametric sensitivity analysis indicates that the key drivers of cost-effectiveness are the tuberculosis case fatality rates, the cohort tuberculosis prevalence, and the SOC screening sensitivity (**Appendix figure 5**).

## Discussion

Benchmarked against estimates of country cost-effectiveness thresholds, this study found that all three TDA-based approaches for diagnosing paediatric tuberculosis in children hospitalised with SAM could be cost-effective compared to SOC from a health systems perspective in Uganda and Zambia. The TB-Speed SAM two-step TDA was cheaper than the TB-Speed SAM one-step and WHO TDAs because its first step resulted in fewer assessments overall. The WHO TDA was dominated by the TB-Speed SAM two-step TDA because it was more costly and less effective. The TB-Speed one-step TDA had the highest costs and health impact. When multiple interventions are compared, the one with the highest expected net benefit is the cost-effective option at a particular cost-effectiveness threshold.^27^ TB-Speed one-step was optimal in Zambia for thresholds over $115/DALY averted, and in Uganda for thresholds over $98/DALY averted. It was also the most effective algorithm. Ultimately, the choice of cost-effectiveness threshold is for local decision makers.

Tuberculosis prevalence was a key determinant of cost-effectiveness: at lower prevalence, only the TB-Speed two-step approach may be cost-effective. This finding has important implications for decentralised use of TDAs in healthcare settings where tuberculosis prevalence is typically lower. The TB-Speed one-step TDA had the highest sensitivity, followed by the TB-Speed two-step TDA. The TDA-based approaches achieved higher sensitivity at the expense of lower specificity, with the WHO TDA having the highest rate of overdiagnosis (43%). Overtreatment is important for resource footprint and inappropriate treatment may negatively impact children and their caregivers. If TDAs are decentralised to lower healthcare levels the number of false positives is likely to be higher. The overall sensitivity of the diagnostic approach for tuberculosis is the key driver of health impact due to the high case fatality rate for children with untreated tuberculosis (61% in the TB-SAM cohort). From the TB-Speed SAM cohort data, we found the case fatality rate was higher in children with SAM and without tuberculosis (12%) than in children with tuberculosis and receiving tuberculosis treatment (9%). Although the difference is small, it may reflect genuinely better outcomes when correctly identifying a serious but treatable disease.

The overall sensitivity of the diagnostic approach for tuberculosis depends not only on the TDA or other assessments, but also on the screening and reassessment steps. Our sensitivity analysis identified the sensitivity of the SOC screening as the third most influential factor for cost-effectiveness. The WHO tuberculosis screening approach includes weight loss >2 weeks, whereas failure to respond to nutritional therapy is suggested as more appropriate for children hospitalised with SAM.^5^ Our sensitivity analyses indicate that weight loss should still be considered for WHO TDA in children with SAM, in order to maintain sensitivity (**Appendix figure 6**). Reassessment was less important for the TDA-based approaches, which had high overall sensitivity. We represented reassessment as full tuberculosis reassessment, including Xpert testing and CXR, and used TB-Speed SAM data to determine the accuracy of these procedures in children previously assessed as non-TB. If data become available on sign and symptom progression, it may become possible to model the use of repeated TDAs in reassessment. In line with our findings, Debes et al. found that case fatality rate for untreated tuberculosis, SOC specifications, and tuberculosis prevalence were major determinants of cost-effectiveness.^28^

Many limitations of our study stem from having less robust data to characterise the SOC arm. For example, we used a screening coverage of 80% for SOC based on expert opinion. For the TDA-based approaches, we assumed complete coverage of systematic tuberculosis screening in children hospitalised with SAM, in line with WHO guidelines and the intended use of the TB-Speed TDAs. Real-life clinical practice may not achieve full screening coverage. Other studies have found screening characteristics are important for cost-effectiveness. Van’t Hoog et al.^29^ explored combinations of sensitivity, specificity and cost at which a hypothetical triage test would improve affordability of the Xpert assay. They found that a triage test with sensitivity equal to Xpert, 75% specificity, and costs of US$5 per patient tested could reduce total diagnostic costs by 42% in the Uganda setting, and by 34% and 39% respectively in the India and South Africa settings. Our sensitivity analyses showed that the SOC and WHO TDA screening specifications were key determinants of cost-effectiveness, thus providing more comprehensive evidence about the impact of a tuberculosis triage test on the overall cost-effectiveness of the TDA-based strategies compared to SOC. For all groups, due to a lack of data on the progression of signs and symptoms, tuberculosis rescreening was not permitted.

Nevertheless, TDA-based approaches demonstrated high screening sensitivity (80% to 100%), indicating that rescreening would likely have minimal impact on these groups. Additional data, however, would be beneficial for the SOC. We also were not able to capture patient costs; more empirical data is needed characterising the economic impact on households affected by paediatric tuberculosis. Lastly, we did not account for a potential negative health impact on children who are inappropriately treated for tuberculosis and costs for their caregivers.

Conversely, our study had major strengths in basing the diagnostic accuracy^14^, clinical outcomes underlying modelling assumptions and also the costs on primary empirical data collection and analysis conducted in the TB-Speed SAM study conducted in two high tuberculosis incidence countries. These assumptions were applied within a framework that accounted for the complexity of patient pathways, including tuberculosis screening and follow-up assessments. Additionally, we developed new unit cost parameters for tuberculosis care specifically for children hospitalised with SAM.

Further research is needed to assess the total health and budget impact of TDA-based interventions, as well as their cost-effectiveness when adapted to different populations of children and implemented in decentralised settings, where availability of tests such as abdominal ultrasound may vary. Future studies should also explore patient costs, the combination of improved child tuberculosis diagnosis strategies with tuberculosis disease severity assessment and eligibility for shortened anti-tuberculosis treatments, which have been shown to be highly cost-effective.^30^ Additionally, the potential of innovative technologies, such as artificial intelligence for reading CXR, should be evaluated to further enhance tuberculosis diagnosis and treatment strategies. Research priorities are outlined in the 2022 WHO consolidated guidelines.^31^

In considering the relevance of these results to other contexts, decision-makers will need to take into account relevant cost-effectiveness thresholds and resources available, the applicability of the case fatality rates and tuberculosis prevalence in the population studied in this analysis, and the relative importance placed on the false positive rate. Different TDAs may be preferred in different contexts, but this study provides strong evidence that any of the three TDAs may be preferred to current care. The WHO has conditionally recommended incorporating TDAs, pending validation, into existing case detection strategies to support the decentralisation of clinical tools and improve the identification of tuberculosis in children. To our knowledge, this is the first study to assess the cost-effectiveness of using treatment decision algorithms in childhood tuberculosis services. This analysis focuses on the vulnerable group of children hospitalised with SAM, and contributes valuable evidence to support the interim WHO recommendation on decentralised models of care. Our findings show that TDA-based approaches are highly cost-effective for the vulnerable group of children hospitalised with SAM, compared to current practices.

## Contributors

MH, MDE, NM, PD, OM, MB, EW conceived the health economics analysis plan. PD developed the cost-effectiveness model. MDE, MH, PD and NM had access to and verified the underlying study data. MDE and MH coordinated international economic data collection and design of the patient pathways. MDE conducted the cost and the cost-effectiveness analyses. MDE, PD, MH, NM interpreted economic results. OM, CC, MB, EW, CR provided scientific expertise and guidance. OM, CC, MB, EW, GB, PS, BN, VM provided clinical expertise.

OM, MB, EW conceived and designed the TB-Speed Severe Acute Malnutrition study, and acquired the project financial support. OM, MB, and EW led the study at international level. EW led the study in Uganda, MHTNN conducted the statistical analysis of the TB-Speed Severe Acute Malnutrition study.

MDE wrote the first draft and all authors reviewed, edited and approved the final version of the manuscript. MDE, PD, OM, CC, MB, EW were responsible for the decision to submit the manuscript.

## Data sharing

Aggregated data for all analyses will be publicly available with the publication on a GitHub repository under a Creative Commons Attribution (CC-BY 4.0) licence (URL: https://github.com/petedodd/TBSsam).

## Data Availability

All data produced are available online at

https://github.com/petedodd/TBSsam

## Acknowledgments

We thank all the children and their families who participated in the study and the healthcare workers of the participating hospitals and laboratories. We thank the Ministries of Health and National Tuberculosis Programmes of participating countries for their support. We thank members of the TB-Speed Scientific Advisory Board who gave technical advice on the design of the study and approved the protocol: Anneke Hesseling (Stellenbosch University, Cape Town, South Africa), Luis Cuevas (Liverpool School of Tropical Medicine, Liverpool, UK), Malgorzata Grzemska and Sabine Verkuijl (WHO, Geneva, Switzerland), Philippa Musoke (Makerere University, Kampala, Uganda), and Mark Nicol (University of Western Australia, Perth, WA, Australia).

