## Appendix for "Treatment decision algorithms for tuberculosis screening and diagnosis in children below 5 years hospitalised with severe acute malnutrition: a cost-effectiveness analysis"

**1. TB-Speed SAM children’s characteristics	2**

Table 1. TB-Speed SAM children’s characteristics overall and by TB status	2

Sampling synthetic cohorts matching feature cross-tabulations	4

**2. Sensitivity and specificity of the each screening approach based on the TB-Speed SAM cohort data	5**

Table 2. Sensitivity and specificity of the each screening approach based on the TB-Speed SAM cohort data	5

**3. TB-Speed SAM and WHO treatment decision algorithm diagrams	6**

Figure 1. TB-Speed SAM one-step treatment decision algorithm3	6

Figure 2. TB-Speed SAM two-step treatment decision algorithm3	7

Figure 3. WHO treatment decision algorithm A (for settings with CXR)2	8

**4. Approaches to tuberculosis reassessment	9**

Table 3. Sensitivity and specificity of each SOC tuberculosis assessment based on the TB-Speed SAM cohort data	11

Table 4.a. Coefficients for generating the sensitivity and specificity of the reassessment exam based on the initial tuberculosis assessment	12

Table 4.b. Example for generating the sensitivity and specificity of the reassessment after clinical assessment only	12

Figure 4. Tuberculosis reassessment as observed in the TB-Speed SAM cohort study	13

**5. Model parameters	14**

Table 5. Cost parameters (2021 US$)	14

Table 6. Costs by activities and by country (2021 US$)	15

Table 7. Non-cost parameters	16

**6. Discounted and undiscounted costs and cost-effectiveness model outputs at varying tuberculosis prevalence, country cost-effectiveness thresholds	18**

Table 8. Costs and cost-effectiveness of the tuberculosis diagnostic approaches by country at varying tuberculosis prevalence	18

Table 9. Country level cost-effectiveness thresholds in cost per DALY averted (2021 US$)	21

**7. Sensitivity analysis of all model parameters on estimated ICERs	22**

Figure 5. Sensitivity analysis of all model parameters (showing only top 10) on estimated ICERs compared to the SOC, by TDA-based approach and by country	24

**8. Sensitivity analysis on the definition of the tuberculosis screening step for the WHO TDA	25**

Figure 6. Effect of the definition of the TB screening step for the WHO TDA on the incremental cost-effectiveness ratios	27

**9. Reference list	28**

### 1. TB-Speed SAM children’s characteristics

#### Table 1. TB-Speed SAM children’s characteristics overall and by TB status

| **Characteristic** | **Overall, N = 535** | **%** | **95% CI** | **TB, N = 101** | **%** | **95% CI** | **NotTB, N = 434** | **%** | **95% CI** |
| --- | --- | --- | --- | --- | --- | --- | --- | --- | --- |
| History of TB contact* | 17 | 3.2 | [1.92; 5.14] | 10 | 9.9 | [5.11; 17.9] | 7 | 1.6 | [0.709; 3.44] |
| Fatigue or loss of playfulness >2 weeks** | 237 | 44.3 | [40.1; 48.6] | 61 | 60.4 | [50.1; 69.8] | 176 | 40.6 | [35.9; 45.4] |
| Fever >2 weeks* & ** | 66 | 12.3 | [9.73; 15.5] | 22 | 21.8 | [14.4; 31.3] | 44 | 10.1 | [7.54; 13.5] |
| Cough >2 weeks* & ** | 96 | 17.9 | [14.8; 21.5] | 31 | 30.7 | [22.1; 40.8] | 65 | 15.0 | [11.8; 18.8] |
| Cough >3 weeks | 63 | 11.8 | [9.23; 14.9] | 25 | 24.8 | [16.9; 34.5] | 38 | 8.8 | [6.35; 11.9] |
| Weight loss >2 weeks** | 333 | 62.2 | [58.0; 66.3] | 67 | 66.3 | [56.2; 75.2] | 266 | 61.3 | [56.5; 65.9] |
| Weight loss | 435 | 81.3 | [77.7; 84.5] | 82 | 81.2 | [71.9; 88.0] | 353 | 81.3 | [77.3; 84.8] |
| Loss of appetite > 2 weeks** | 250 | 46.7 | [42.4; 51.1] | 63 | 62.4 | [52.1; 71.7] | 187 | 43.1 | [38.4; 47.9] |
| Temperature > 38 °C | 24 | 4.5 | [2.96; 6.70] | 14 | 13.9 | [8.06; 22.5] | 10 | 2.3 | [1.18; 4.34] |
| Tachycardia | 61 | 11.4 | [8.90; 14.5] | 23 | 22.8 | [15.3; 32.4] | 38 | 8.8 | [6.35; 11.9] |
| Tachypnea | 20 | 3.7 | [2.36; 5.81] | 9 | 8.9 | [4.41; 16.7] | 11 | 2.5 | [1.34; 4.63] |
| Cervical or supra-clavicular adenopathy | 8 | 1.5 | [0.697; 3.04] | 4 | 4.0 | [1.28; 10.4] | 4 | 0.9 | [0.296; 2.51] |
| Chest-in-drawing | 42 | 7.9 | [5.78; 10.5] | 19 | 18.8 | [12.0; 28.1] | 23 | 5.3 | [3.46; 7.96] |
| Crackles on auscultation | 73 | 13.6 | [10.9; 16.9] | 29 | 28.7 | [20.4; 38.7] | 44 | 10.1 | [7.54; 13.5] |
| Depressed level of consciousness | 269 | 50.3 | [46.0; 54.6] | 62 | 61.4 | [51.1; 70.8] | 207 | 47.7 | [42.9; 52.5] |
| CXR - Presence of miliary | 6 | 1.1 | [0.457; 2.55] | 4 | 4.0 | [1.28; 10.4] | 2 | 0.5 | [0.080; 1.84] |
| CXR - Presence of alveolar opacity | 134 | 25.0 | [21.5; 29.0] | 45 | 44.6 | [34.8; 54.8] | 89 | 20.5 | [16.9; 24.7] |
| CXR - Presence of hilar/mediastinal lymphadenopathy | 146 | 27.3 | [23.6; 31.3] | 45 | 44.6 | [34.8; 54.8] | 101 | 23.3 | [19.4; 27.6] |
| CXR - Presence of excavation (cavity) | 7 | 1.3 | [0.575; 2.80] | 2 | 2.0 | [0.344; 7.66] | 5 | 1.2 | [0.425; 2.83] |
| CXR - Presence of pleural effusion | 7 | 1.3 | [0.575; 2.80] | 5 | 5.0 | [1.84; 11.7] | 2 | 0.5 | [0.080; 1.84] |
| CXR - Presence of pericardial effusion | 6 | 1.1 | [0.457; 2.55] | 1 | 1.0 | [0.052; 6.18] | 5 | 1.2 | [0.425; 2.83] |
| AUS Splenic micro abscesses | 25 | 4.7 | [3.11; 6.92] | 15 | 14.9 | [8.82; 23.6] | 10 | 2.3 | [1.18; 4.34] |
| AUS Hepatic micro abscesses | 4 | 0.7 | [0.240; 2.04] | 2 | 2.0 | [0.344; 7.66] | 2 | 0.5 | [0.080; 1.84] |
| AUS Pericardial or pleural effusion | 72 | 13.5 | [10.7; 16.7] | 23 | 22.8 | [15.3; 32.4] | 49 | 11.3 | [8.55; 14.7] |
| AUS Peritoneal effusion - ascites | 43 | 8.0 | [5.94; 10.8] | 11 | 10.9 | [5.83; 19.0] | 32 | 7.4 | [5.17; 10.4] |
| Xpert result positive | 43 | 8.0 | [5.94; 10.8] | 43 | 42.6 | [32.9; 52.8] | 0 | 0.0 | [0.000; 1.09] |
| HIV positive | 58 | 10.8 | [8.40; 13.9] | 19 | 18.8 | [12.0; 28.1] | 39 | 9.0 | [6.54; 12.2] |

*used for the screening step of the SOC, **used for the screening step of the WHO TDA

#### Sampling synthetic cohorts matching feature cross-tabulations

To be able to sample synthetic cohorts with the right frequency of signs/symptoms (features) and correlations between them, we used cross-tabulations of feature–pair counts for children designated by the expert committee as having tuberculosis or not. We used the ‘mvdc’ function from the R package copula (version 1.1-4) to fit normal copulas with beta marginals to this data. Synthetic cohorts for children with and without tuberculosis were then sampled. TB-Speed SAM data lacked information on haemoptysis and night sweats. These features were independently sampled by tuberculosis status based on the frequency data in Gunasekera et al. (used to develop the WHO TDA), and added to the synthetic cohorts.^1^

### 2. Sensitivity and specificity of each screening approach based on the TB-Speed SAM cohort data

#### Table 2. Sensitivity and specificity of each screening approach based on the TB-Speed SAM cohort data

| **Screening approach (presence of any of the signs/symptoms)** | **TB,**  **n = 101** | **Sensitivity** | | **NotTB,**  **n = 434** | **Specificity** | |
| --- | --- | --- | --- | --- | --- | --- |
|  |  | **%** | **95% CI** |  | **%** | **95% CI** |
| **SOC**: Screening 2 (Fever >2 weeks, Cough >2 weeks, Contact TB) | 40 | 37.0 | [28.5-46.4] | 393 | 79.4 | [75.6-82.7] |
| **TB-Speed One-step:** No screening (all children considered presumptive TB) | 101 | 100 |  | 0 | 0 |  |
| **TB-Speed Two-step:** Screening 5 (History of TB contact, Cough >3 weeks, Temperature >38°C, Tachycardia, Chest-in-drawing, Crackles on auscultation, Depressed level of consciousness, Cervical or supra-clavicular adenopathy, HIV positive) | 95 | 88 | [80.5-92.8] | 170 | 34.3 | [30.3-38.6] |
| **WHO-TDA:** Screening* (Fever >2 weeks, Cough >2 weeks, Fatigue or loss of playfulness >2 weeks, Loss of appetite >2 weeks, Weight loss >2 weeks) | 85 | 78.7 | [70.0-85.4] | 139 | 28.1 | [24.3-32.2] |

*as recommended in the 2022 WHO operational handbook for childhood TB^2^

### 3. TB-Speed SAM and WHO treatment decision algorithm diagrams

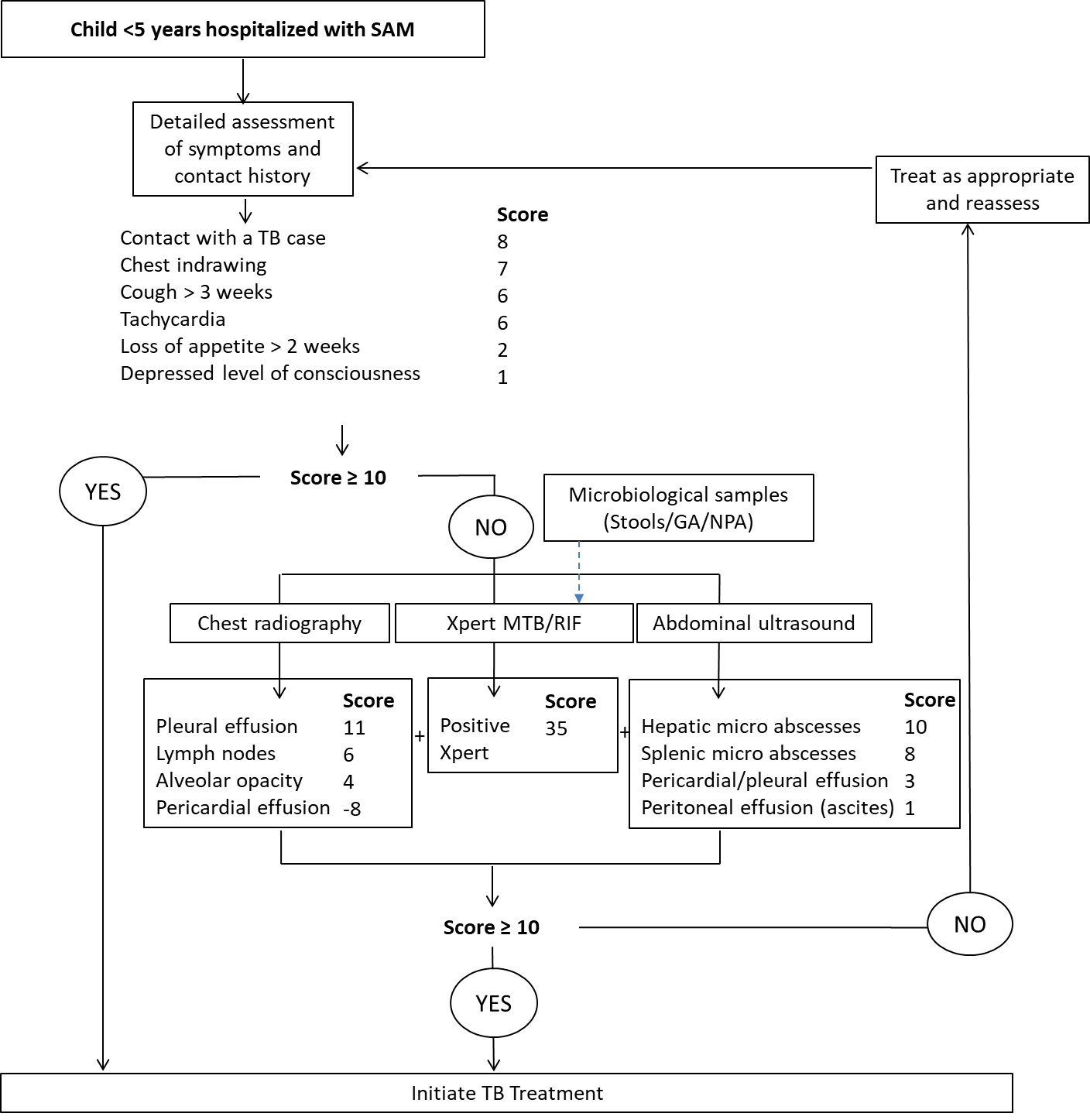

#### Figure 1. TB-Speed SAM one-step treatment decision algorithm^3^

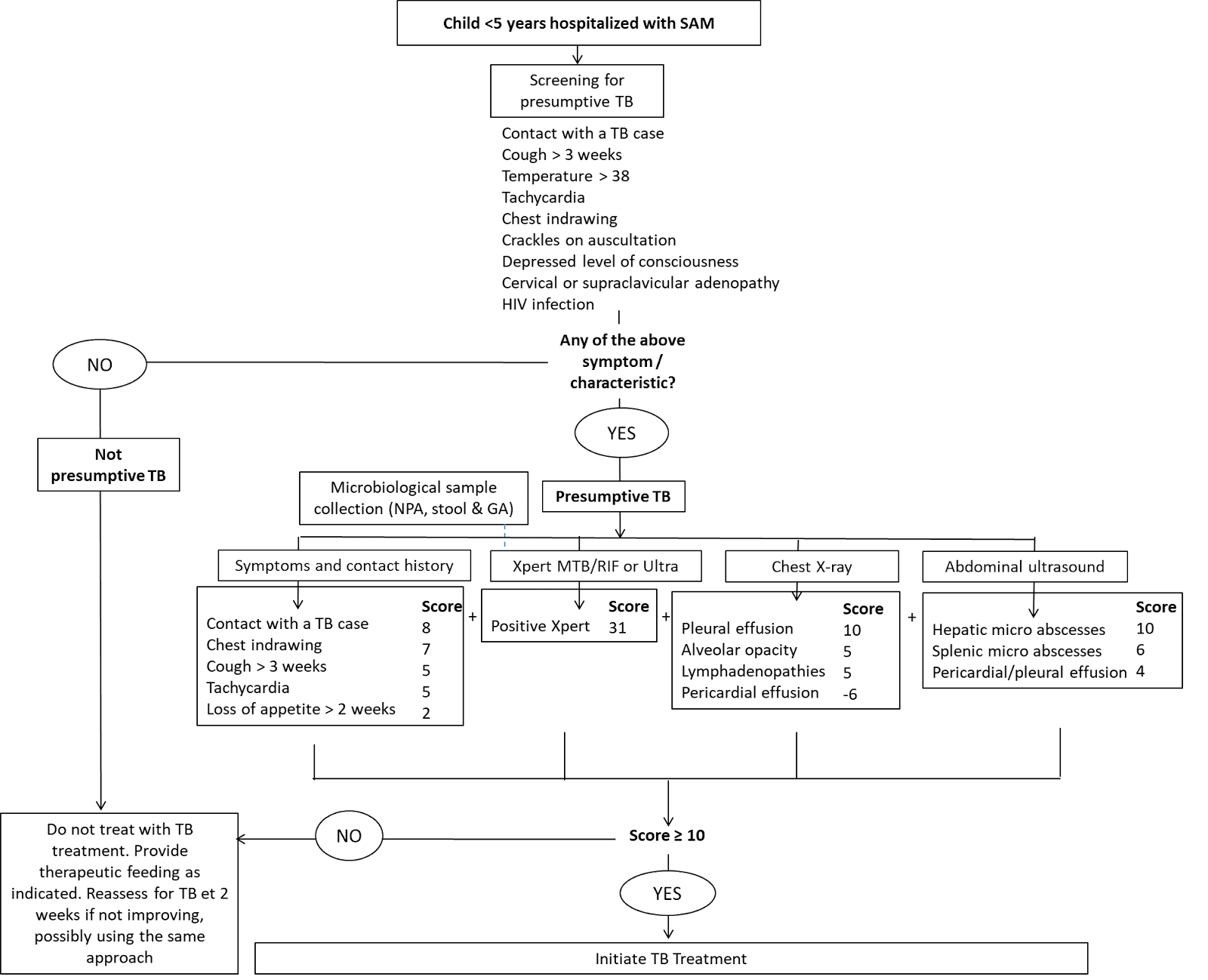

#### Figure 2. TB-Speed SAM two-step treatment decision algorithm^3^

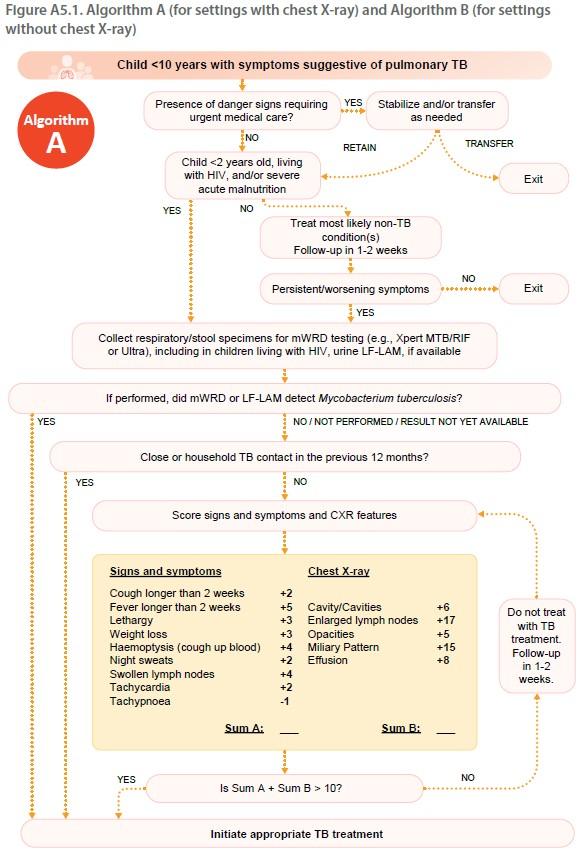

#### Figure 3. WHO treatment decision algorithm A (for settings with CXR)^2^

### 4. Approaches to tuberculosis reassessment

Using the TB-Speed SAM cohort data (**Figure 4**), we defined a two-step reassessment process using:

1. The clinician’s choice to reassess where the sensitivity is: true TB who are reassessed / true TB not diagnosed at the initial clinical assessment, and specificity: true non-TB not reassessed / true non-TB not diagnosed at the initial clinical assessment,
2. Reassessment exam’s sensitivity and specificity

Reassessment was taken to comprise CXR, Xpert on gastric aspirate, and clinical assessment, and their sensitivity and specificity were conditional on results of (clinical) components used in initial assessment being negative.

a. *Standard of care arm*

Let ${N^{+}}_{i}$ be those with true TB who are positive to test combination *i ,* and ${N^{-}}_{i}$ those with truly TB negative who are test negative to test combination *i*. Let $N^{\pm}$ be the total number true positive/negative, respectively. Combinations of tests are assumed to be positive if any of the tests are positive. This means the specificities multiply:

$$Sp_{123}=Sp_{1}\times Sp_{2}\times Sp_{3}$$

And 1 − sensitivities (call them $\bar{Se}=1-Se)$ multiply:

$$\bar{Se}_{123}=\bar{Se}_{1}\times\bar{Se}_{2}\times\bar{Se}_{3}$$

We are interested in understanding the sensitivity of combinations of tests conditional on a negative result initially. CXR and most of the features defining clinical assessment are not likely to be very dynamic between initial and reassessment. We are assuming that reassessment comprises clinical assessment, Xpert on GA, and CXR, and that reassessment after initial negative assessment on this combination will remain the same. As an example, to calculate the sensitivity and specificity of reassessment after an initial assessment that was only clinical assessment (and which was negative, i.e. unit specificity), we are interested in:

$$Sp_{reassess|clin}=Sp_{clin,CXR,XGA}=(Sp_{clin}=1)\times Sp_{CXR}\times Sp_{XGA}=Sp_{CXR}\times Sp_{XGA}=\frac{Sp_{clin,CXR,XGA}}{Sp_{clin}}$$

And similarly

$\bar{Se}_{reassess|clin}=\frac{\bar{Se}_{clin,CXR,XGA}}{\bar{Se}_{clin}}$

We have data to define the point estimates of sensitivity and specificities in these numerators and denominators,

$$Sp_{reassess|clin}=\frac{{N^{-}}_{clin,CXR,XGA}}{{N^{-}}_{clin}}$$

$$\bar{Se}_{reassess|clin}=\frac{1-{N^{+}}_{clin,CXR,XGA}/N^{+}}{1-{N^{+}}_{clin}/N^{+}}$$

but wish to parametrise beta distribution. To do this we used corresponding numerator denominator counts and assume a uniform prior for a conjugate analysis, so that:

$$\bar{Se}_{reassess|clin}\sim B\left( 1+{{N^{+}-N}^{+}}_{clin,CXR,XGA},1+{N^{+}}_{clin,CXR,XGA}{{-N}^{+}}_{clin} \right)$$

$$Sp_{reassess|clin}\sim B\left( 1+{N^{-}}_{clin,CXR,XGA},1+{N^{-}}_{clin}-{N^{-}}_{clin,CXR,XGA} \right)$$

*Example for estimating the sensitivity and specificity of reassessment after clinical assessment only:*

From **table 3**, we extract the following coefficients into **tables 4.a.** and **4.b.**

#### Table 3. Sensitivity and specificity of each SOC tuberculosis assessment based on the TB-Speed SAM cohort data

| **SOC TB exams, among those screened + in SOC** (Fever >2 weeks OR Cough >2 weeks OR Contact TB) | **TB,**  **n = 40** | **Sensitivity** | | **NotTB,**  **n = 102** | **Specificity** | |
| --- | --- | --- | --- | --- | --- | --- |
|  |  | **%** | **95% CI** |  | **%** | **95% CI** |
| **clinical exam only:**  cough> 2 weeks OR  contact TB | 36 | 90 | [75-97] | 26 | 25 | [18-35] |
| **clinical exam + CXR:**  cough> 2 weeks OR  contact TB OR  CXR - Presence of miliary OR  CXR - Presence of hilar/mediastinal lymphadenopathy OR  CXR - Presence of excavation) | 39 | 98 | [85-100] | 14 | 14 | [8-23] |
| **clinical exam + Xpert:**  cough> 2 weeks OR  contact TB OR  Xpert(GA) | 37 | 92 | [79-98] | 25 | 25 | [17-35] |
| **clinical exam + CXR + Xpert (GA):**  cough> 2 weeks OR  contact TB OR  CXR - Presence of miliary OR  CXR - Presence of hilar/mediastinal lymphadenopathy OR  CXR - Presence of excavation) OR  Xpert(GA) | 40 | 100 | [89-100] | 14 | 14 | [8-23] |

#### Table 4.a. Coefficients for generating the sensitivity and specificity of the reassessment exam based on the initial tuberculosis assessment

| **Parameter N by TB status and type of exam** | **Number of children** |
| --- | --- |
| $N^{+}$ | 40* |
| $N^{-}$ | 102 |
| ${N^{+}}_{clin,CXR,XGA}$ | 40* |
| ${N^{-}}_{clin,CXR,XGA}$ | 14* |
| ${N^{+}}_{clin}$ | 36* |
| ${N^{-}}_{clin}$ | 26* |
| ${N^{+}}_{clin,CXR}$ | 39 |
| ${N^{-}}_{clin,CXR}$ | 14 |
| ${N^{+}}_{clin,XGA}$ | 37 |
| ${N^{-}}_{clin,XGA}$ | 25 |

*used in the example

$$\bar{Se}_{reassess|clin}\sim B\left( 1+40-40,1+40-36 \right) =B\left( 1,5 \right)$$

$$Sp_{reassess|clin}\sim B\left( 1+14,1+26-14 \right)=B(15,13)$$

#### Table 4.b. Example for generating the sensitivity and specificity of the reassessment after clinical assessment only

| **NAME** | **DISTRIBUTION** | **MEDIAN (IQR)** | **DESCRIPTION** |
| --- | --- | --- | --- |
| s.soc.reassessafterclin.sebar | B(1,5) | 0.129 (0.056 - 0.242) | SOC: 1 − sensitivity of reassessment after clinical assessment only |
| s.soc.reassessafterclin.sp | B(15,13) | 0.537 (0.472 - 0.600) | SOC: specificity of reassessment after clinical assessment only |

b. *TDA-based approaches*

*Step 1:*

Sensitivity of choice to reassess = TP/(TP+FN) = (27 + 9 + 5)/(27 + 9 + 5 + 4) = 41/45 = 91%

Specificity of choice to reassess = TN/(TN+FP) = 355/(355 + 9 + 125) = 355/489= 73%

*Step 2:*

Sensitivity of reassessment exam = 27/(27+9+5) = 27/41 = 65%

Specificity of reassessment exam = 125/(125+9) = 125/134 = 93%

*Note*: We considered that TB found post-mortem was classified as not diagnosed.

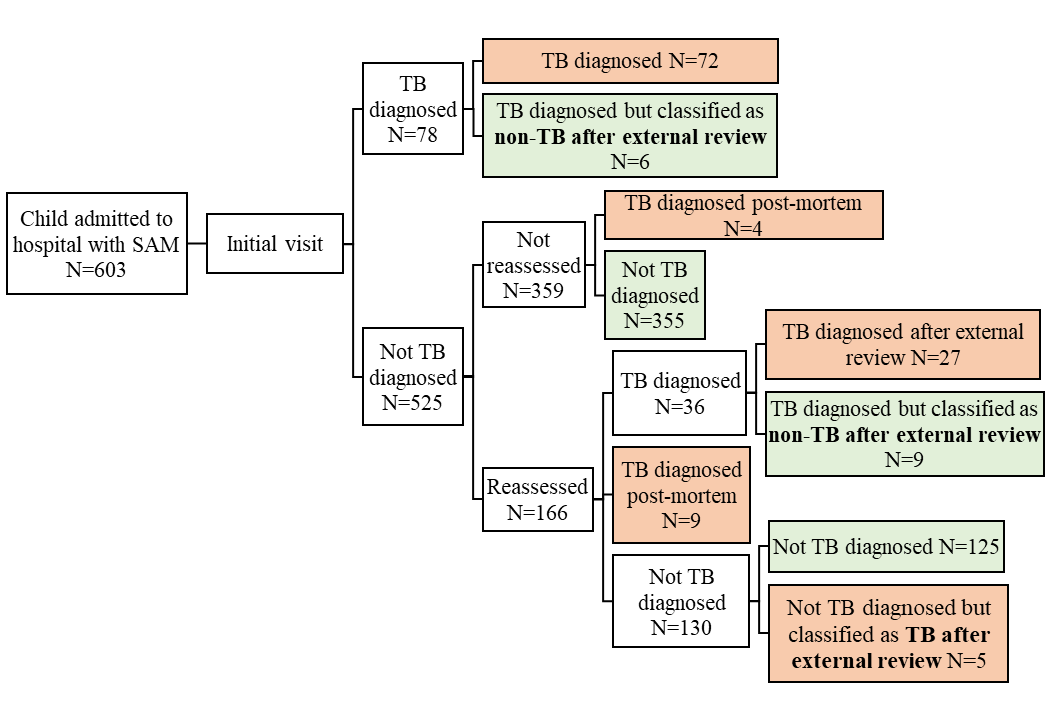

#### Figure 4. Tuberculosis reassessment as observed in the TB-Speed SAM cohort study

### 5. Model parameters

#### Table 5. Cost parameters (2021 US$)

| **NAME** | **DESCRIPTION** | **Uganda** | | **Zambia** | |
| --- | --- | --- | --- | --- | --- |
|  |  | **Low** | **High** | **Low** | **High** |
| c.s.soc.scre | SOC: TB screening | 0.28 | 0.33 | 0.21 | 0.28 |
| c.s.soc.exam | SOC: initial clinical assessment | 2.78 | 3.61 | 3.88 | 4.58 |
| c.s.soc.CXR | SOC: CXR [for all] | 7.13 | 8.00 | 7.76 | 8.39 |
| c.s.soc.CXRxga | SOC: CXR [for all] + Xpert Ultra on GA [for all] + Urine LF-LAM | 52.79 | 65.10 | 61.06 | 73.70 |
| c.s.soc.xga | SOC: Xpert Ultra on GA [for all] + Urine LF-LAM | 45.66 | 57.10 | 53.30 | 65.32 |
| c.s.tbs1step.diag.clin | INT: TB-Speed one-step algo: clinical exam + CXR + Xpert on GA and stool | 69.10 | 79.24 | 79.16 | 89.91 |
| c.s.tbs1step.diag | INT: TB-Speed one-step algo: clinical exam + CXR + Xpert on NPA and stool + abdo US | 72.74 | 84.98 | 84.80 | 97.10 |
| c.s.tbs2step.scre | INT: TB-Speed two-steps algo: screening: clinical exam + HIV test | 6.65 | 7.77 | 7.64 | 8.67 |
| c.s.tbs2step.diag | INT: TB-Speed two-steps algo: diagnostic: CXR + Xpert on NPA and stool + abdo US | 69.96 | 81.37 | 80.93 | 92.53 |
| c.s.who.scre | INT: WHO algo: TB screening | 0.28 | 0.33 | 0.21 | 0.28 |
| c.s.who.hiv.diag | INT: WHO algo: clinical exam + TB contact + CXR + Xpert on NPA and stool + HIV test + Urine LF-LAM | 82.12 | 98.29 | 92.07 | 108.89 |
| c.s.who.diag | INT: WHO algo: clinical exam + TB contact + CXR + Xpert on NPA and stool + HIV test | 72.97 | 83.40 | 82.92 | 94.00 |
| c.s.rsATT | Rifampicin-sensitive anti-TB treatment in SAM children | 34.69 | 40.67 | 43.21 | 47.79 |
| c.s.rrATT | Rifampicin-resistant anti-TB treatment in SAM children | 1458.94 | 1506.09 | 1526.15 | 1562.24 |
| c.s.reassessCXRxgastall | Reassessment exam + CXR [for all] + Xpert Ultra on GA and stool [for all] | 67.64 | 77.63 | 77.70 | 88.28 |

#### Table 6. Costs by activities and by country (2021 US$)

|  | **Uganda** | | **Zambia** | |
| --- | --- | --- | --- | --- |
| **Activity details** | **Low** | **High** | **Low** | **High** |
| Systematic TB screening | 0.28 | 0.33 | 0.21 | 0.28 |
| 1. Initial clinical examination: history, symptoms, physical exam; or  2. Second check-up at 7 days | 1.32 | 2.00 | 2.42 | 2.95 |
| Initial clinical examination: vital signs, measurements | 1.46 | 1.61 | 1.46 | 1.63 |
| Blood test - HIV | 3.87 | 4.16 | 3.76 | 4.09 |
| Urine LAM test | 9.15 | 14.89 | 9.15 | 14.89 |
| Collecting sputum sample | 1.17 | 1.18 | 1.19 | 1.23 |
| Collecting GA sample | 15.85 | 18.92 | 23.01 | 26.38 |
| Collecting NPA sample | 11.51 | 14.40 | 13.27 | 16.22 |
| Collecting (and preparing) stool sample | 2.02 | 2.14 | 2.23 | 2.48 |
| Processing sample (NPA, stool, GA, sputum) and conducting Xpert Ultra test | 20.66 | 23.28 | 21.14 | 24.05 |
| Smear microscopy - all sample types - Lab resources | 0.73 | 0.74 | 0.83 | 0.92 |
| Conducting digital chest X-ray | 7.13 | 8.00 | 7.76 | 8.39 |
| Conducting abdominal ultrasound | 3.64 | 5.74 | 5.64 | 7.19 |
| Diagnosis & Treatment - DS-TB treatment: children 0-4 years with SAM | 34.69 | 40.67 | 43.21 | 47.79 |
| Diagnosis & Treatment - MDR-TB treatment : children 0-4 years with SAM | 1458.94 | 1506.09 | 1526.15 | 1562.24 |

#### Table 7. Non-cost parameters

| **NAME** | **DISTRIBUTION** | **MEDIAN (IQR)** | **DESCRIPTION** | **SOURCE** |
| --- | --- | --- | --- | --- |
| s.soc.testingcov | B(30.21696,3.35744) | 0.908 (0.871 - 0.938) | SOC: coverage of any testing among children with presumptive TB (the remaining children only receive clinical assessment) | Expert opinion |
| s.soc.CXRonly | B(47.52,47.52) | 0.500 (0.465 - 0.535) | SOC: fraction of those with tests only having CXR (as opposed Xpert Ultra on GA as well) | Expert opinion |
| s.soc.Xpertonly | B(17.7575, 53.2725) | 0.248 (0.214 - 0.283) | SOC: fraction of those with tests only having Xpert Ultra on GA (as opposed CXR as well) | Expert opinion |
| s.TBprev | B(101,434) | 0.188 (0.177 - 0.200) | True TB prevalence (committee definition) | TB-Speed SAM cohort data |
| s.reassess.choice.sp | B(356,135) | 0.733 (0.719 - 0.746) | Specificity of choice to reassess | TB-Speed SAM cohort data |
| s.reassess.choice.se | B(42,5) | 0.899 (0.867 - 0.926) | Sensitivity of choice to reassess | TB-Speed SAM cohort data |
| s.soc.scrcov | B(48.37248,12.09312) | 0.800 (0.700 - 0.900) | Proportion of children being screened for TB when admitted to hospital with SAM | Expert opinion |
| s.soc.screen.se | B(40.99281,69.79857) | 0.370 (0.285 - 0.464) | Sensitivity of the SOC screening | TB-Speed SAM cohort data |
| s.soc.screen.sp | B(395.0861,102.5034) | 0.794 (0.782 - 0.806) | Specificity of the SOC screening | TB-Speed SAM cohort data |
| s.CFR.sam.noTB | B(60,437) | 0.120 (0.111 - 0.130) | CFR of children with SAM but not TB, with or without ATT | TB-Speed SAM cohort data |
| s.CFR.sam.TBATT | B(9,83) | 0.095 (0.076 - 0.117) | CFR of children with SAMand TB, with ATT | TB-Speed SAM cohort data |
| s.CFR.sam.TBnoATT | B(12,8) | 0.603 (0.528 - 0.676) | CFR of children with SAMand TB, without ATT | TB-Speed SAM cohort data |
| s.soc.clin.se | B(37,5) | 0.887 (0.851 - 0.917) | SOC: sensitivity of clinical assessment | TB-Speed SAM cohort data |
| s.soc.clin.sp | B(27,77) | 0.258 (0.230 - 0.288) | SOC: specificity of clinical assessment | TB-Speed SAM cohort data |
| s.soc.clinCXR.se | B(40,2) | 0.959 (0.936 - 0.977) | SOC: sensitivity of clinical assessment with CXR | TB-Speed SAM cohort data |
| s.soc.clinCXR.sp | B(15,89) | 0.142 (0.120 - 0.166) | SOC: specificity of clinical assessment with CXR | TB-Speed SAM cohort data |
| s.soc.clinGA.se | B(38,4) | 0.911 (0.879 - 0.938) | SOC: sensitivity of clinical assessment with Xpert on GA | TB-Speed SAM cohort data |
| s.soc.clinGA.sp | B(26,78) | 0.248 (0.221 - 0.278) | SOC: specificity of clinical assessment with Xpert on GA | TB-Speed SAM cohort data |
| s.soc.clinCXRGA.se | B(41,1) | 0.983 (0.967 - 0.993) | SOC: sensitivity of clinical assessment with CXR & Xpert on GA | TB-Speed SAM cohort data |
| s.soc.clinCXRGA.sp | B(15,89) | 0.142 (0.120 - 0.166) | SOC: specificity of clinical assessment with CXR & Xpert on GA | TB-Speed SAM cohort data |
| s.soc.reassessafterclin.sebar | B(1,5) | 0.129 (0.056 - 0.242) | SOC: 1 − sensitivity of reassessment after clinical assessment only | TB-Speed SAM cohort data |
| s.soc.reassessafterclin.sp | B(15,13) | 0.537 (0.472 - 0.600) | SOC: specificity of reassessment after clinical assessment only | TB-Speed SAM cohort data |
| s.soc.reassessafterclinCXR.sebar | B(1,2) | 0.293 (0.134 - 0.500) | SOC: 1 − sensitivity of reassessment after clinical assessment with CXR | TB-Speed SAM cohort data |
| s.soc.reassessafterclinCXR.sp | B(15,1) | 0.955 (0.912 - 0.981) | SOC: specificity of reassessment after clinical assessment with CXR | TB-Speed SAM cohort data |
| s.soc.reassessafterclinGA.sebar | B(1,4) | 0.159 (0.069 - 0.293) | SOC: 1 − sensitivity of reassessment after clinical assessment with Xpert on GA | TB-Speed SAM cohort data |
| s.soc.reassessafterclinGA.sp | B(15,12) | 0.557 (0.491 - 0.621) | SOC: specificity of reassessment after clinical assessment with Xpert on GA | TB-Speed SAM cohort data |
| s.nonsocreassess.sp | B(126,10) | 0.929 (0.913 - 0.942) | Non-SOC: specificity of reassessment | TB-Speed SAM cohort data |
| s.nonsocreassess.se | B(28,15) | 0.654 (0.603 - 0.702) | Non-SOC: sensitivity of reassessment | TB-Speed SAM cohort data |

### 6. Discounted and undiscounted costs and cost-effectiveness model outputs at varying tuberculosis prevalence, country cost-effectiveness thresholds

#### Table 8. Costs and cost-effectiveness of the tuberculosis diagnostic approaches by country at varying tuberculosis prevalence

|  | **Arm (vs. Comparator)** | **Uganda** | **Zambia** | **Uganda** | **Zambia** | **Uganda** | **Zambia** |
| --- | --- | --- | --- | --- | --- | --- | --- |
| **Tuberculosis prevalence (%)** | | **18.9%^a^** | | **10.0%** | | **5.0%** | |
| **Per child hospitalised with SAM** | | | | | | | |
| Costs per child (95% UI) in 2021 US dollars | SOC - Total | 32 (29 to 36) | 38 (33 to 42) | 32 (28 to 36) | 37 (32 to 41) | 31 (28 to 35) | 36 (32 to 41) |
|  | WHO TDA - Total | 85 (79 to 92) | 98 (90 to 105) | 83 (77 to 90) | 95 (88 to 103) | 82 (76 to 89) | 94 (87 to 102) |
|  | Two-step TDA - Total | 82 (75 to 88) | 94 (87 to 102) | 78 (71 to 86) | 89 (82 to 99) | 76 (69 to 83) | 87 (79 to 96) |
|  | One-step TDA - Total | 108 (104 to 112) | 125 (121 to 129) | 106 (102 to 110) | 122 (118 to 128) | 105 (101 to 109) | 121 (117 to 126) |
| Incremental costs per child (95% UI) in 2021 US dollars | WHO TDA (vs. SOC) | 53 (46 to 60) | 60 (52 to 68) | 52 (44 to 59) | 59 (50 to 67) | 51 (44 to 58) | 58 (49 to 66) |
|  | Two-step TDA (vs. SOC) | 49 (42 to 57) | 57 (48 to 65) | 46 (39 to 54) | 53 (44 to 62) | 44 (37 to 52) | 51 (42 to 60) |
|  | One-step TDA (vs. SOC) | 75 (70 to 80) | 87 (81 to 93) | 74 (69 to 79) | 86 (80 to 91) | 73 (68 to 79) | 85 (79 to 91) |
|  | Two-step TDA (vs WHO TDA) | -3 (-11 to 4) | -3 (-12 to 5) | -6 (-13 to 2) | -6 (-15 to 3) | -7 (-15 to 1) | -7 (-16 to 2) |
|  | One-step TDA (vs. WHO TDA) | 23 (16 to 29) | 27 (20 to 35) | 22 (16 to 29) | 27 (19 to 34) | 22 (16 to 29) | 27 (20 to 35) |
|  | One-step TDA (vs. Two-step TDA) | 26 (20 to 32) | 31 (24 to 37) | 28 (22 to 34) | 33 (26 to 39) | 29 (23 to 35) | 34 (27 to 41) |
| **Per 100 children hospitalised with SAM** | | | | | | | |
| Deaths (95% UI) | SOC - Total | 19 (16 to 21) | 19 (16 to 21) | 16 (13 to 20) | 16 (13 to 20) | 14 (12 to 18) | 14 (12 to 18) |
|  | WHO TDA - Total | 14 (13 to 15) | 14 (13 to 15) | 13 (12 to 15) | 13 (12 to 15) | 13 (12 to 14) | 13 (12 to 14) |
|  | Two-step TDA - Total | 13 (12 to 15) | 13 (12 to 14) | 13 (12 to 14) | 13 (12 to 14) | 12 (12 to 13) | 12 (12 to 14) |
|  | One-step TDA - Total | 12 (12 to 13) | 12 (12 to 13) | 12 (12 to 13) | 12 (12 to 13) | 12 (12 to 13) | 12 (12 to 13) |
| Deaths averted (95% UI) | WHO TDA (vs. SOC) | 5 (3 to 7) | 5 (3 to 7) | 3 (0 to 6) | 3 (0 to 6) | 1 (0 to 5) | 1 (0 to 5) |
|  | Two-step TDA (vs. SOC) | 5 (3 to 7) | 5 (3 to 8) | 3 (0 to 7) | 3 (0 to 7) | 1 (0 to 5) | 1 (0 to 5) |
|  | One-step TDA (vs. SOC) | 6 (4 to 9) | 6 (4 to 8) | 3 (1 to 8) | 3 (1 to 8) | 2 (0 to 6) | 2 (0 to 6) |
|  | Two-step TDA (vs WHO TDA) | 0 (-1 to 2) | 0 (-1 to 2) | 0 (-1 to 1) | 0 (0 to 1) | 0 (0 to 1) | 0 (0 to 1) |
|  | One-step TDA (vs. WHO TDA) | 1 (0 to 3) | 1 (0 to 3) | 1 (0 to 2) | 1 (0 to 2) | 0 (0 to 2) | 0 (0 to 2) |
|  | One-step TDA (vs. Two-step TDA) | 1 (0 to 2) | 1 (0 to 2) | 0 (0 to 1) | 0 (0 to 1) | 0 (0 to 1) | 0 (0 to 1) |
| Discounted DALYs (95% UI) | SOC - Total | 509 (449 to 570) | 509 (450 to 571) | 424 (346 to 552) | 424 (347 to 547) | 378 (329 to 502) | 378 (330 to 504) |
|  | WHO TDA - Total | 377 (347 to 414) | 378 (347 to 415) | 355 (329 to 396) | 355 (329 to 397) | 343 (326 to 381) | 343 (326 to 382) |
|  | Two-step TDA - Total | 364 (337 to 396) | 364 (337 to 394) | 347 (327 to 382) | 347 (328 to 380) | 339 (325 to 367) | 339 (326 to 369) |
|  | One-step TDA - Total | 338 (321 to 358) | 338 (321 to 359) | 334 (322 to 352) | 334 (322 to 352) | 332 (324 to 346) | 332 (323 to 347) |
| Incremental discounted DALYs averted (95% UI) | WHO TDA (vs. SOC) | 131 (78 to 190) | 131 (77 to 191) | 69 (9 to 164) | 69 (9 to 167) | 35 (-1 to 129) | 35 (-1 to 126) |
|  | Two-step TDA (vs. SOC) | 145 (88 to 201) | 145 (91 to 205) | 77 (12 to 178) | 77 (12 to 183) | 40 (0 to 141) | 40 (0 to 138) |
|  | One-step TDA (vs. SOC) | 171 (113 to 235) | 171 (113 to 232) | 90 (16 to 206) | 90 (17 to 216) | 47 (0 to 161) | 46 (0 to 160) |
|  | Two-step TDA (vs WHO TDA) | 13 (-20 to 49) | 14 (-20 to 48) | 8 (-15 to 37) | 8 (-13 to 39) | 4 (-12 to 28) | 4 (-11 to 29) |
|  | One-step TDA (vs. WHO TDA) | 40 (10 to 75) | 40 (11 to 76) | 21 (-4 to 60) | 21 (-3 to 58) | 11 (-5 to 45) | 11 (-4 to 48) |
|  | One-step TDA (vs. Two-step TDA) | 26 (7 to 51) | 26 (7 to 51) | 14 (0 to 40) | 13 (0 to 39) | 7 (0 to 29) | 7 (0 to 30) |
| Undiscounted DALYs (95% UI) | SOC - Total | 1247 (1102 to 1398) | 1245 (1100 to 1396) | 1039 (849 to 1354) | 1038 (850 to 1339) | 928 (807 to 1231) | 925 (808 to 1232) |
|  | WHO TDA - Total | 925 (851 to 1014) | 924 (850 to 1016) | 871 (807 to 971) | 869 (805 to 971) | 841 (800 to 934) | 839 (797 to 935) |
|  | Two-step TDA - Total | 892 (827 to 971) | 891 (823 to 964) | 852 (802 to 936) | 849 (801 to 930) | 831 (798 to 900) | 829 (796 to 902) |
|  | One-step TDA - Total | 827 (786 to 877) | 826 (785 to 878) | 819 (790 to 863) | 817 (787 to 862) | 814 (793 to 847) | 812 (790 to 848) |
| Incremental undiscounted DALYs averted (95% UI) | WHO TDA (vs. SOC) | 322 (190 to 465) | 321 (187 to 466) | 169 (22 to 401) | 169 (23 to 408) | 87 (-2 to 317) | 86 (-3 to 308) |
|  | Two-step TDA (vs. SOC) | 355 (215 to 494) | 354 (223 to 501) | 188 (30 to 437) | 188 (29 to 448) | 97 (0 to 346) | 97 (0 to 339) |
|  | One-step TDA (vs. SOC) | 419 (277 to 575) | 418 (276 to 567) | 221 (40 to 506) | 221 (41 to 527) | 114 (0 to 395) | 114 (0 to 391) |
|  | Two-step TDA (vs WHO TDA) | 33 (-50 to 119) | 33 (-49 to 117) | 19 (-36 to 91) | 19 (-33 to 95) | 10 (-28 to 69) | 10 (-27 to 71) |
|  | One-step TDA (vs. WHO TDA) | 98 (24 to 185) | 98 (26 to 187) | 52 (-9 to 147) | 52 (-7 to 142) | 27 (-11 to 111) | 27 (-10 to 118) |
|  | One-step TDA (vs. Two-step TDA) | 65 (18 to 125) | 65 (17 to 126) | 33 (0 to 98) | 33 (0 to 96) | 17 (0 to 72) | 17 (0 to 73) |
| Discounted ICER | WHO TDA (vs. SOC) | 40 | 46 | 75 | 85 | 144 | 163 |
|  | Two-step TDA (vs. SOC) | 34 | 39 | 60 | 68 | 112 | 128 |
|  | One-step TDA (vs. SOC) | 44 | 51 | 82 | 95 | 158 | 182 |
|  | Two-step TDA (vs WHO TDA) | Two-step TDA dominates WHO TDA | Two-step TDA dominates WHO TDA | Two-step TDA dominates WHO TDA | Two-step TDA dominates WHO TDA | Two-step TDA dominates WHO TDA | Two-step TDA dominates WHO TDA |
|  | One-step TDA (vs. Two-step TDA) | 98 | 115 | 207 | 245 | 419 | 492 |
| Undiscounted ICER | WHO TDA (vs. SOC) | 16 | 19 | 31 | 35 | 59 | 67 |
|  | Two-step TDA (vs. SOC) | 14 | 16 | 25 | 28 | 46 | 52 |
|  | One-step TDA (vs. SOC) | 18 | 21 | 34 | 39 | 64 | 74 |
|  | Two-step TDA (vs WHO TDA) | Two-step TDA dominates WHO TDA | Two-step TDA dominates WHO TDA | Two-step TDA dominates WHO TDA | Two-step TDA dominates WHO TDA | Two-step TDA dominates WHO TDA | Two-step TDA dominates WHO TDA |
|  | One-step TDA (vs. Two-step TDA) | 40 | 47 | 85 | 100 | 171 | 201 |

^a^ Prevalence of tuberculosis in the TB-Speed SAM study

#### Table 9. Country level cost-effectiveness thresholds in cost per DALY averted (2021 US$)

|  | **Country** | **Threshold (Ochalek, 2018)** | | **GDP per capita*** |
| --- | --- | --- | --- | --- |
|  |  | **Low** | **High** |  |
| Threshold (% GDP per capita) | Uganda | 150 (17 %) | 194 (22 %) | 884 |
|  | Zambia | 364 (32 %) | 500 (44 %) | 1,137 |

* given for reference, not as a suggested threshold option

### 7. Sensitivity analysis of all model parameters on estimated ICERs

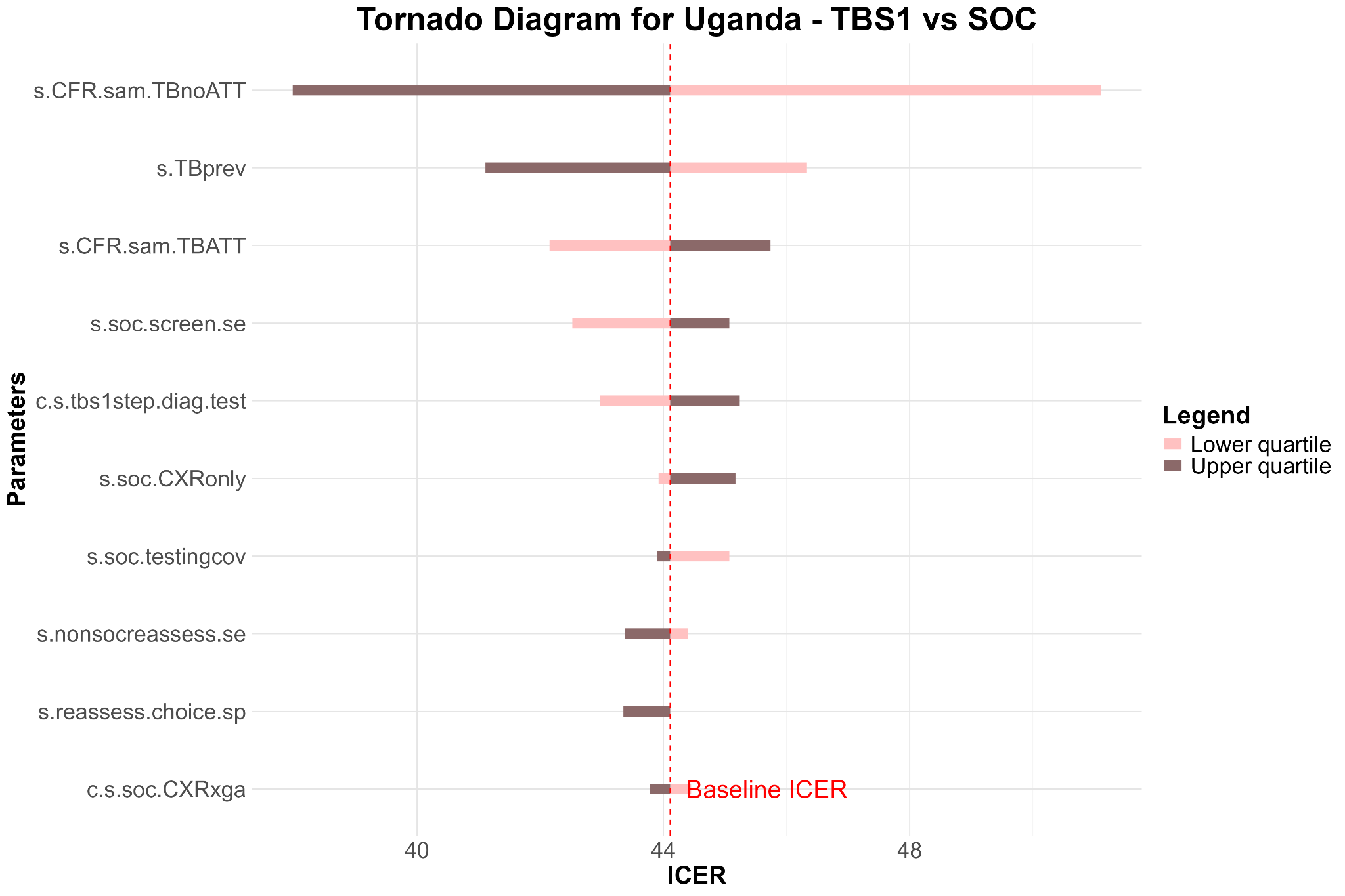

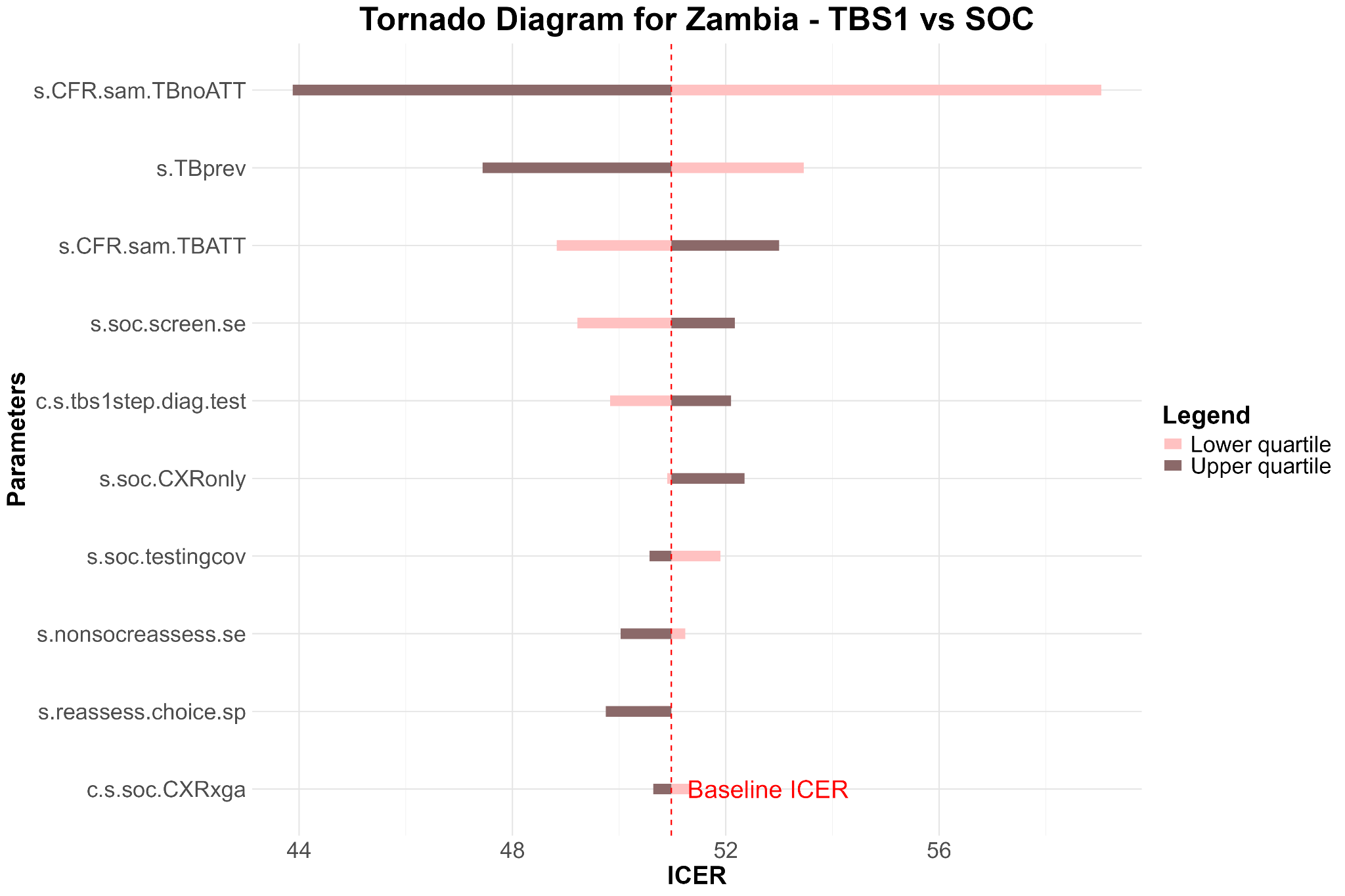

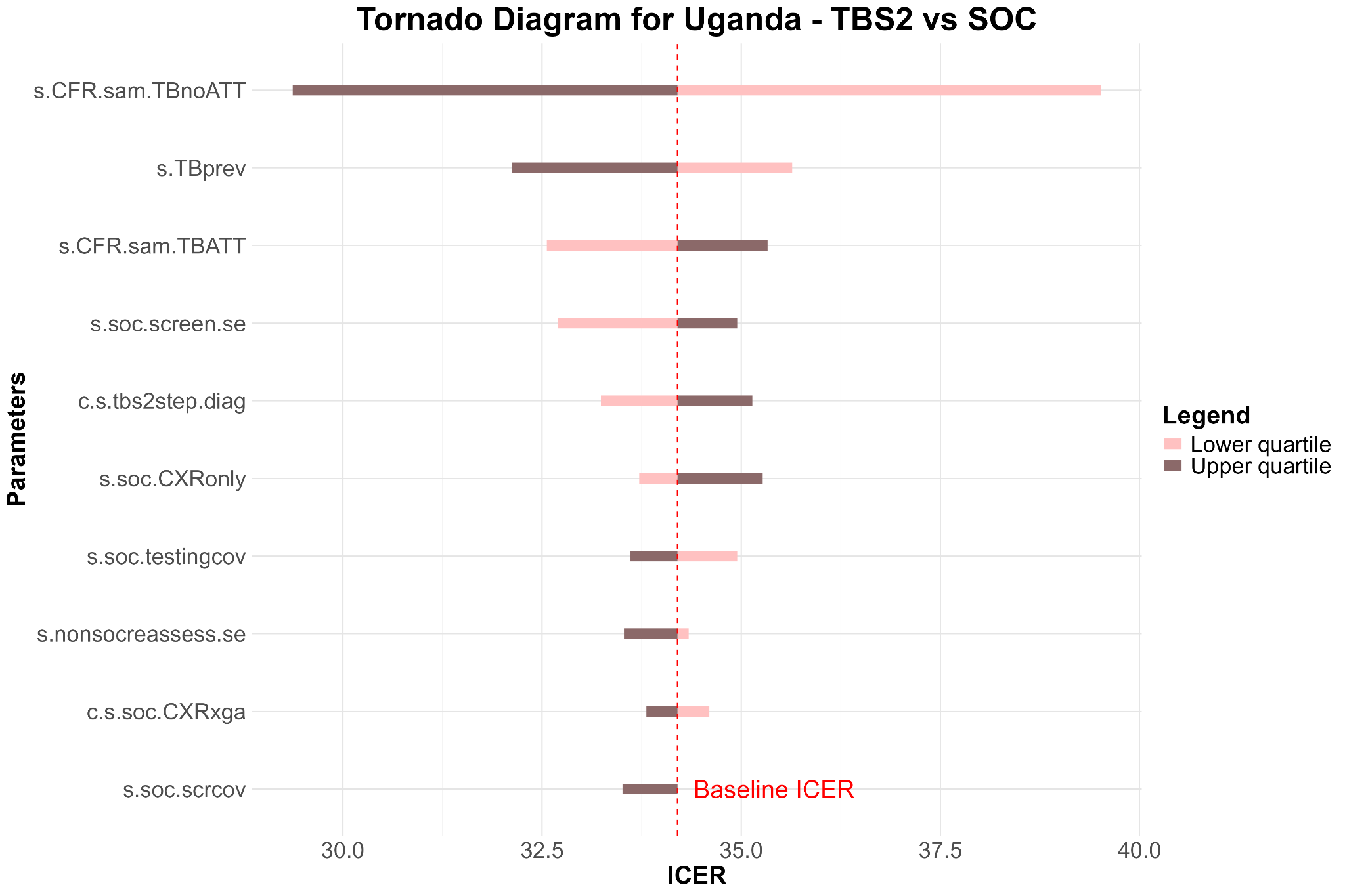

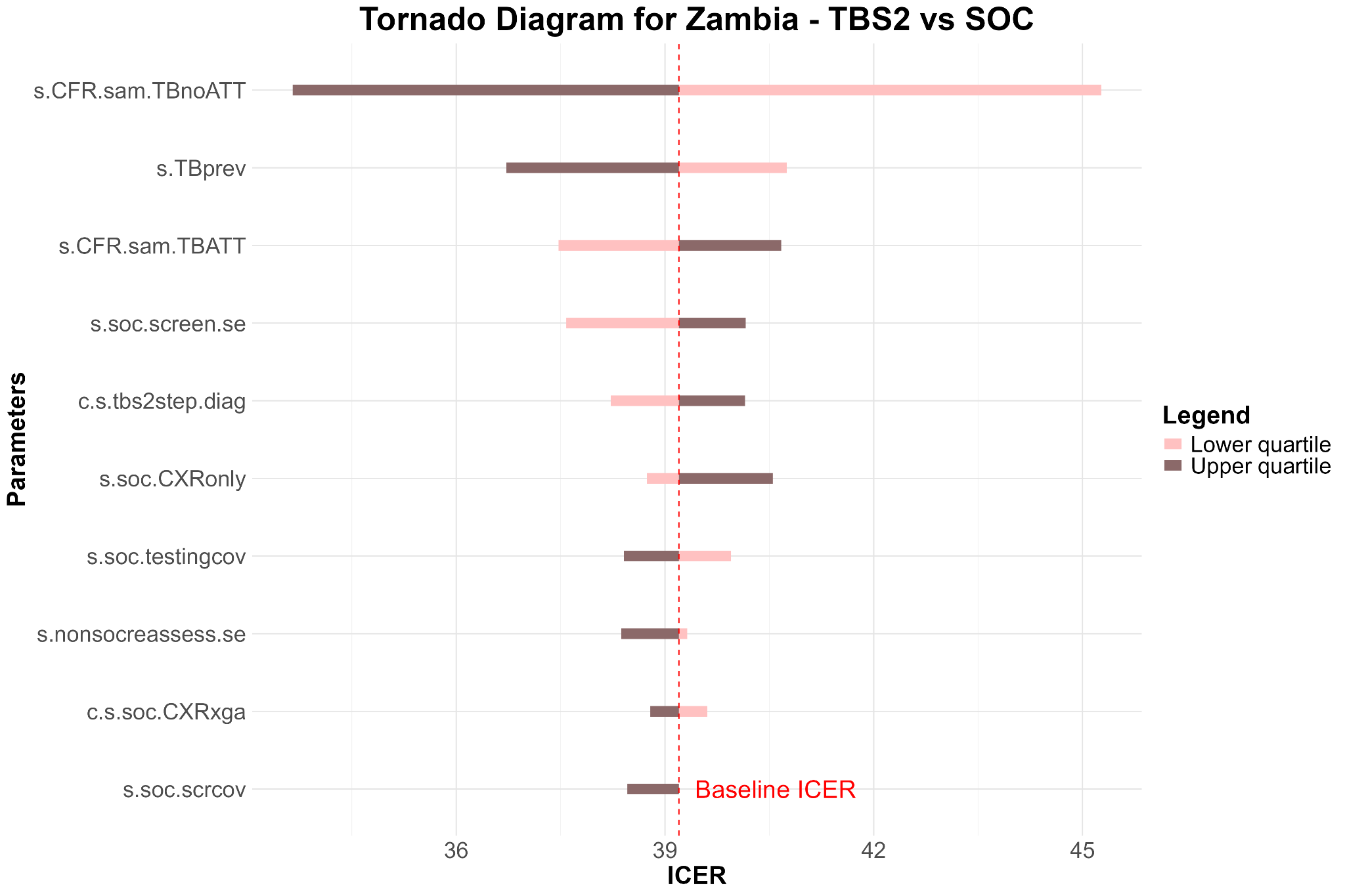

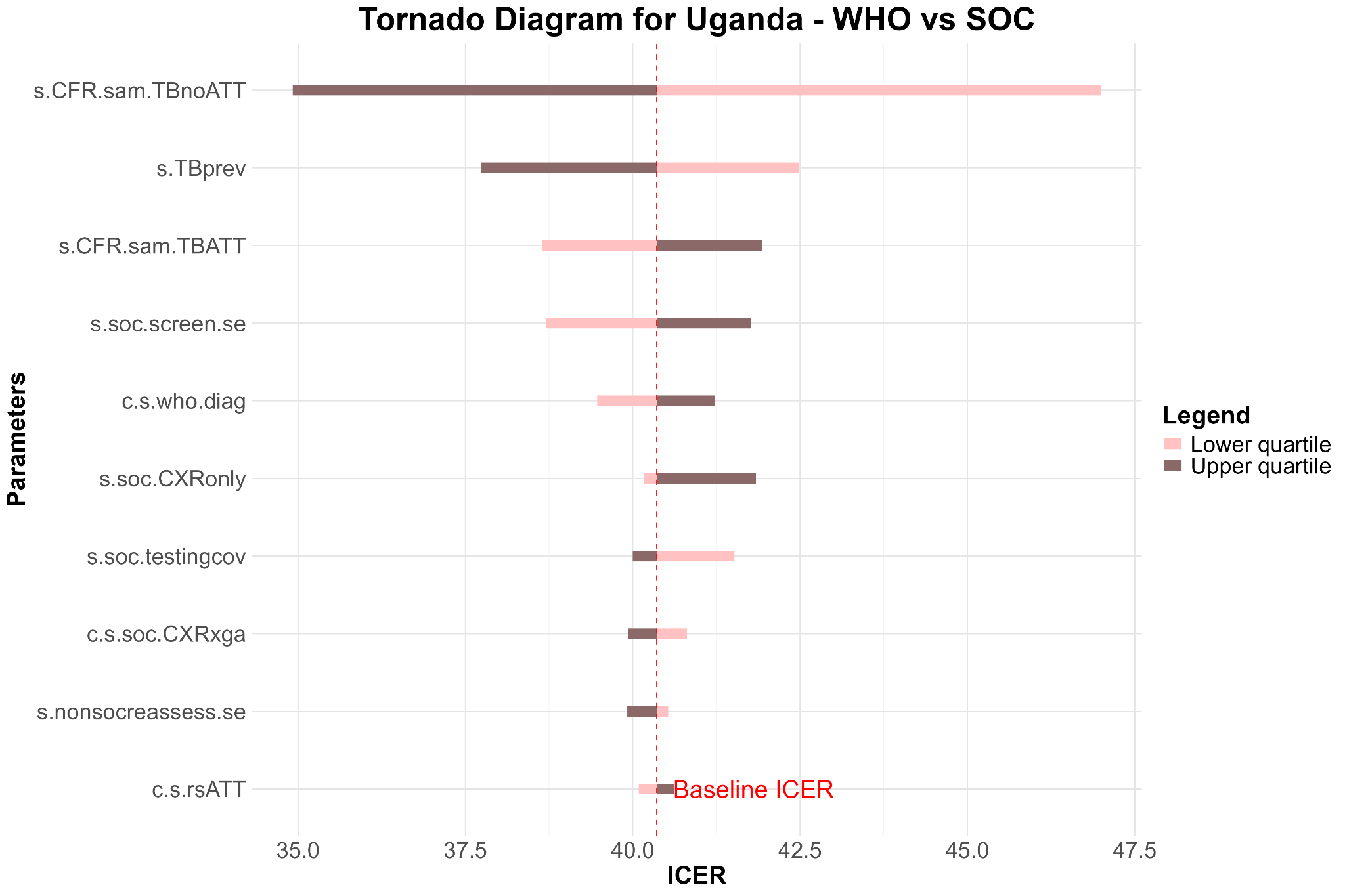

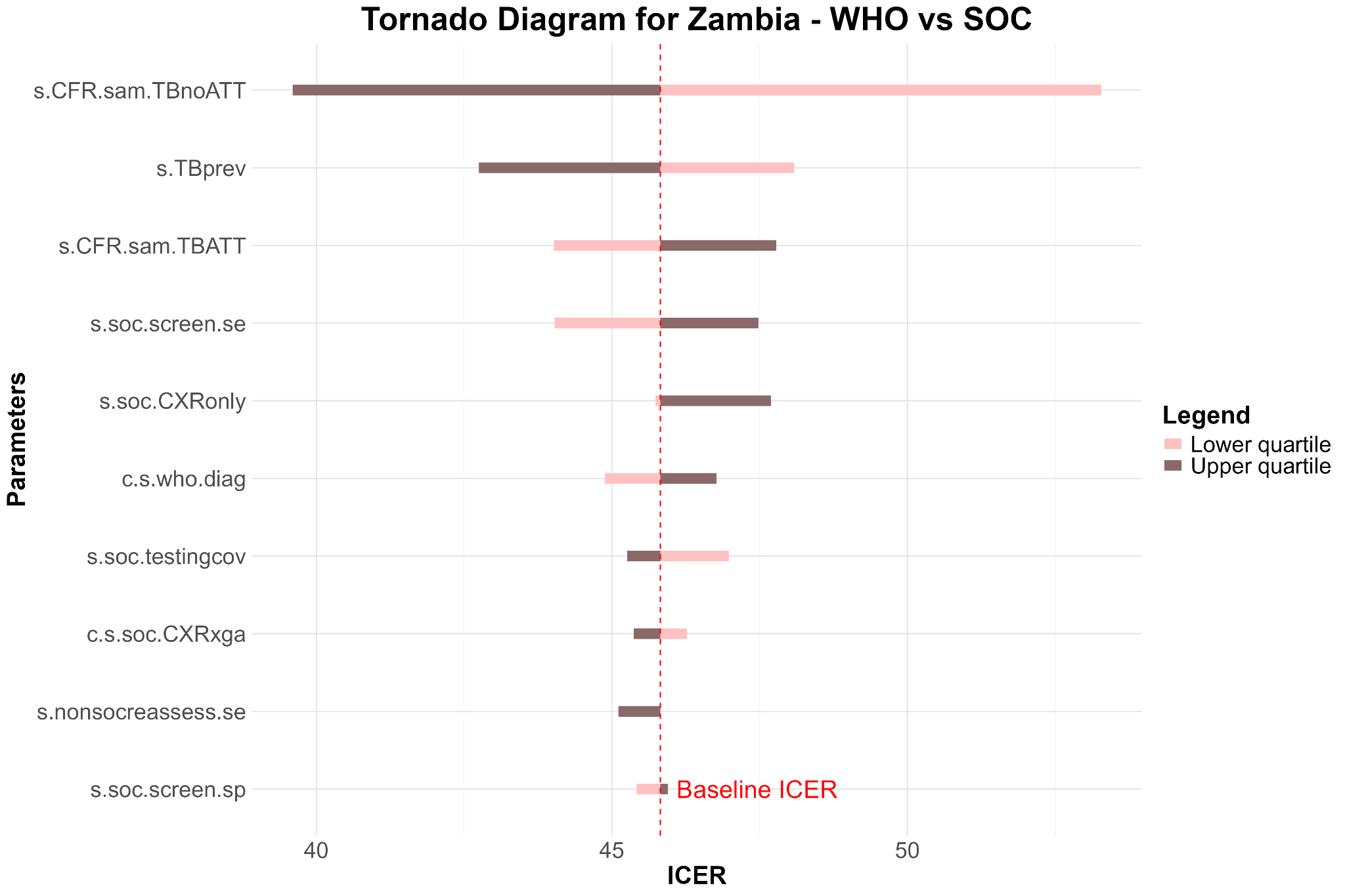

#### Figure 5. Sensitivity analysis of all model parameters (showing only top 10) on estimated ICERs compared to the SOC, by TDA-based approach and by country

*Note*: Segments aligned in the same direction result from rounding errors for parameters with minimal estimated impact on ICERs.

### 8. Sensitivity analysis on the definition of the tuberculosis screening step for the WHO TDA

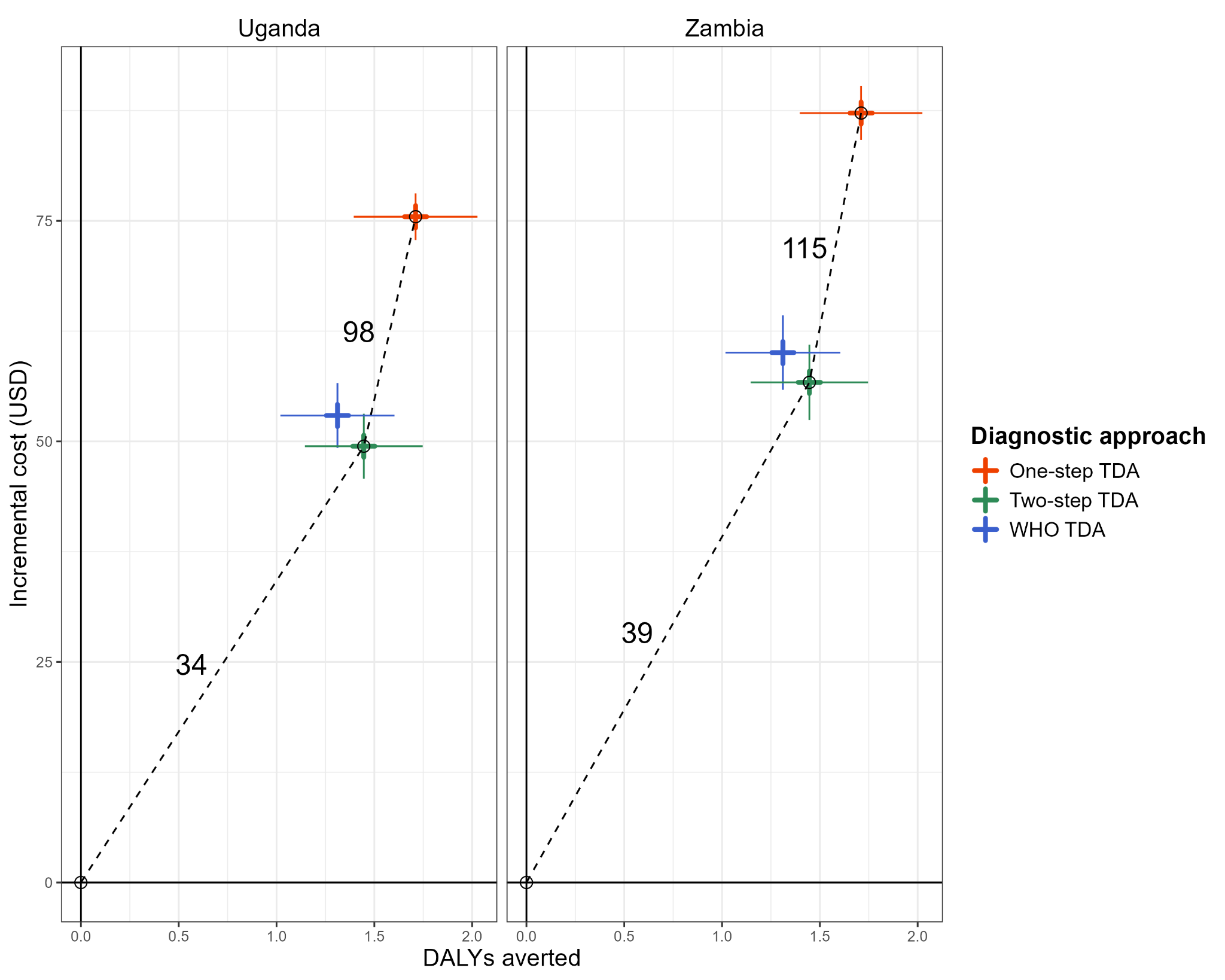

Presence of any of **5** chronic symptoms (>2 weeks): fever, cough, fatigue or loss of playfulness, *weight loss*, and loss of appetite (**base case**)

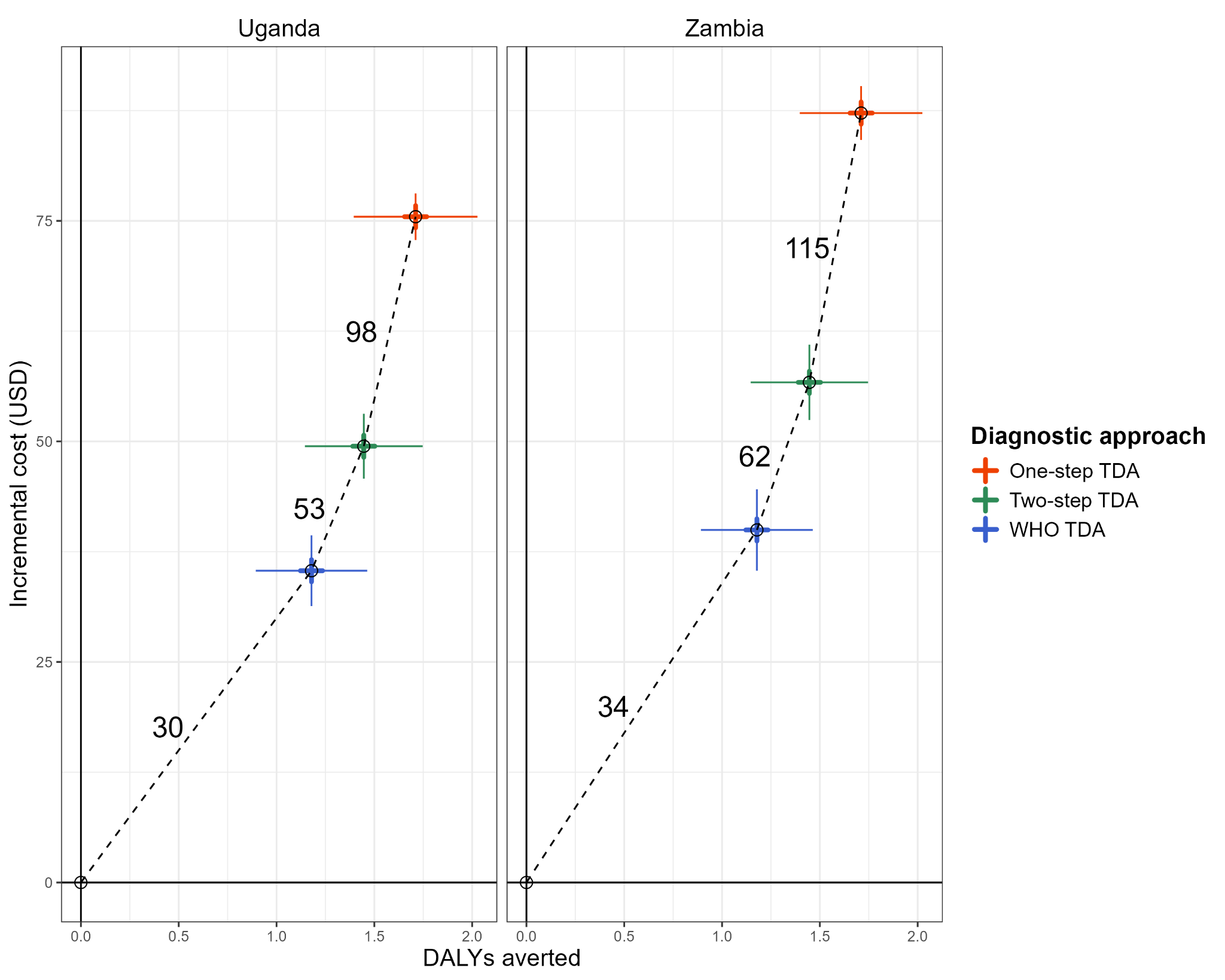

Presence of any of **4** chronic symptoms (>2 weeks): fever, cough, fatigue or loss of playfulness, and loss of appetite

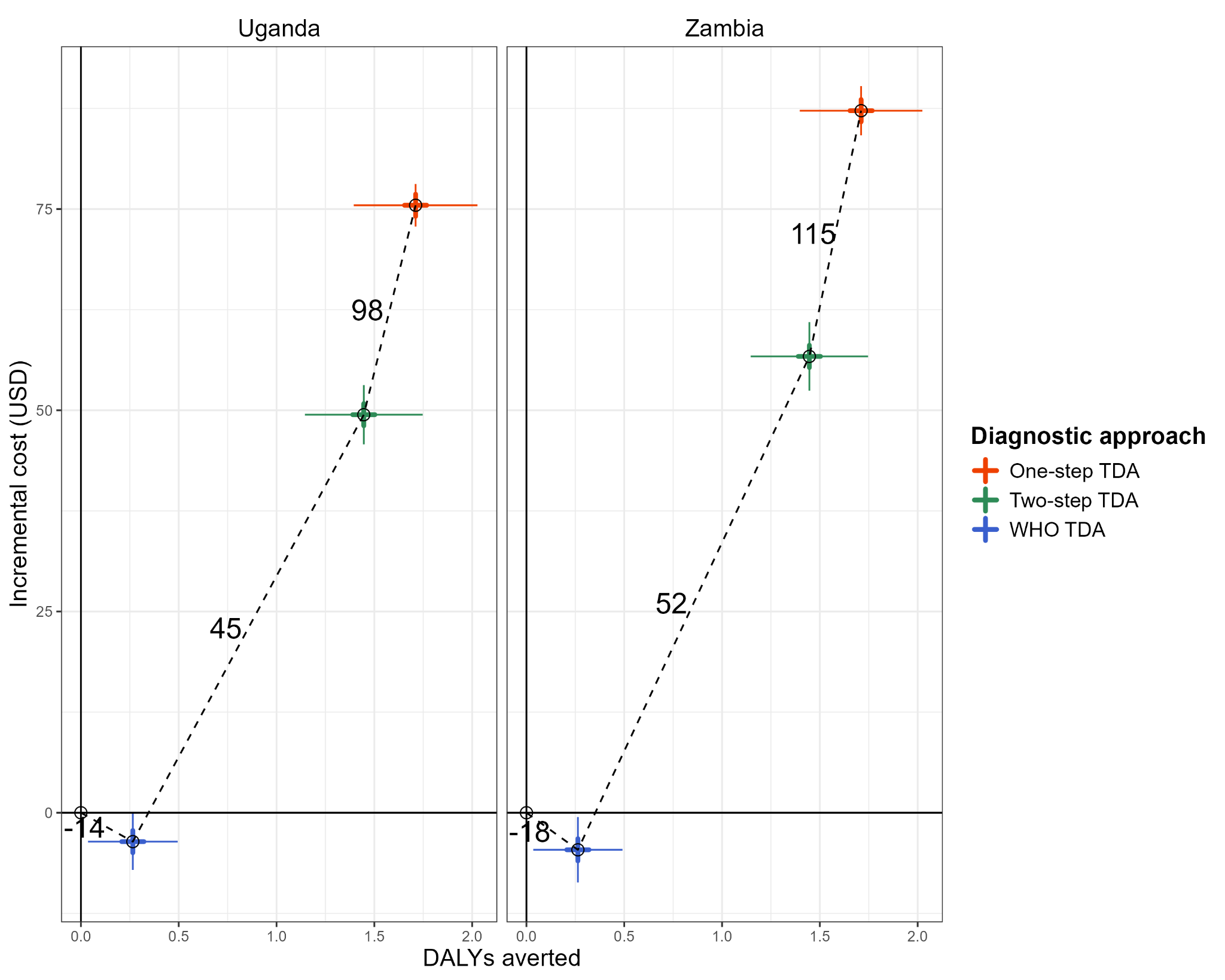

Presence of any of **2** chronic symptoms (>2 weeks): fever, cough, + contact TB history (**as in SOC**)

#### Figure 6. Effect of the definition of the TB screening step for the WHO TDA on the incremental cost-effectiveness ratios
